# Machine learning analysis of a digital insole versus clinical standard gait assessments for digital endpoint development

**DOI:** 10.1101/2022.10.05.22280750

**Authors:** Matthew F. Wipperman, Allen Z. Lin, Kaitlyn M. Gayvert, Benjamin Lahner, Selin Somersan-Karakaya, Xuefang Wu, Joseph Im, Minji Lee, Bharatkumar Koyani, Ian Setliff, Malika Thakur, Daoyu Duan, Aurora Breazna, Fang Wang, Wei Keat Lim, Gabor Halasz, Jacek Urbanek, Yamini Patel, Gurinder S. Atwal, Jennifer D. Hamilton, Clotilde Huyghues-Despointes, Oren Levy, Andreja Avbersek, Rinol Alaj, Sara C. Hamon, Olivier Harari

## Abstract

Biomechanical gait analysis informs clinical practice and research by linking characteristics of gait with neurological or musculoskeletal injury or disease. However, there are limitations to analyses conducted at gait labs as they require onerous construction of force plates into laboratories mimicking the lived environment, on-site patient assessments, as well as requiring specialist technicians to operate. Digital insoles may offer patient-centric solutions to these challenges. In this work, we demonstrate how a digital insole measuring osteoarthritis-specific gait signatures yields similar results to the clinical gait-lab standard. To achieve this, we constructed a machine learning model, trained on force plate data collected in participants with knee arthropathy and healthy controls. This model was highly predictive of force plate data from a validation set (area under the receiver operating characteristics curve (auROC) = 0.86; area under the precision-recall curve (auPR) = 0.90) and of a separate, independent digital insole dataset containing control and knee osteoarthritis subjects (auROC = 0.83; auPR = 0.86). After showing that digital insole derived gait characteristics are comparable to traditional gait measurements, we next show that a single stride of raw sensor time series data could be accurately assigned to each subject, highlighting that individuals (even healthy) using digital insoles can be identified by their gait characteristics. This work provides a framework for a promising alternative to traditional clinical gait analysis methods, adds to the growing body of knowledge regarding wearable technology analytical pipelines, and supports clinical development of at-home gait assessments, with the potential to improve the ease, frequency, and depth of patient monitoring.

**One Sentence Summary:** Biosensor data collected by digital insoles is comparable to lab-based clinical assessments and can be used to identify subject-specific gait patterns.

## Introduction

Gait assessment plays several roles in clinical practice and research for neurological and musculoskeletal diseases: diagnostic workup; guiding treatment selection and measuring response; and assessment of gait and balance pathophysiology.^1^ Traditionally, gait is assessed in the clinic under the supervision of a technician, physical therapist, or research scientist in a specialized gait laboratory with a force platform, with or without a motion tracking system. Gait labs use equipment that enables the creation of extensive models of human movement. These may include force plates to measure ground reaction force (GRF), video analysis to map an individual’s skeletal architecture, and electromyography to measure muscle activity during movement.

A critical module of the gait lab, the force plate, produces electrical signals that can be processed into three components of force (vertical, anterior-posterior, and medio-lateral), as well as the derived characteristic, center of pressure (COP) in the x/y direction. These signals provide information on gait characteristics, postural stability as well as direction, strength, and duration of stance phase.^2,3^ These can provide insight into a patient’s neuromuscular function and guide diagnosis for disorders such as Parkinson’s disease^4,5^ or progressive supranuclear palsy;^6^ disease progression and severity as shown with multiple sclerosis^7^ and osteoarthritis (OA) patients;^8^ and can identify patients with elevated fall risk by examining gait variability and balance.^9^ Drawbacks of force plates that limit their use include cost and space requirements together with the need for trained personnel to interpret the results.

Wearable devices that assess certain gait characteristics in controlled and free-living environments have gained popularity in both commercial and research settings to overcome these challenges. These devices have the potential to bring some aspects of the gait lab into the patient’s home.^10^ A major goal of gait measurements with wearable digital health technologies (DHTs) is to provide information that is comparable to a gait lab assessment in a more accessible, user friendly, remote, and patient-centric manner. In this work, we evaluate Moticon digital insoles, designed for optimization of training in high-performance athletes (Moticon Rego AG, Munich, Germany). Each Moticon digital insole has a total of 25 sensors per foot: 16 vertical plantar pressure sensors that assess force, a trial-axial accelerometer that measures acceleration, and a gyroscope that measures orientation and angular velocity. Each sensor captures data at 100 Hz, and dedicated software computes several clinically relevant spatial and temporal derived gait characteristics comparable to data generated in a gait lab. The Moticon digital insole computes GRF in the same way as a force plate, generating comparable data outputs. In addition to GRF data, derived gait characteristics summarizing a subject’s walk and raw sensor time series data can be obtained. Collectively, the data generated from such digital insoles may provide rich gait phenotyping information to characterize gait in patients with a broad range of neurological and musculoskeletal diseases.

In addition to challenges with collection of data, the use of wearable devices in clinical research presents analytical challenges. Clinical research analysis pipelines should aim to improve diagnosis and monitoring of treatment responses, as well as support the development of meaningful digital endpoints. However, to achieve this, analytical challenges must be solved. Gait quantitation generates dense raw sensor time series data with non-linear relationships that make analysis and interpretation challenging. As a class, wearable sensor data analysis pipelines suffer from a lack of well-established analytical methods compared to other biomarker data types.^11-13^ The novelty of DHT technologies create opportunities to not only establish criterion validity by demonstrating how a digital biomarker replicates a clinical standard, but to further identify digital biomarkers with face validity that accurately measure the concept it purports to, as well as construct validity, which improve upon readouts of existing constructs.

To support digital endpoint development, a machine learning (ML) framework may be used as a tool to evaluate the consistency of data generated from DHTs, as well as how well these data can be used as clinical trial endpoints. Selection of the appropriate modeling modalities for a particular clinical question is challenging.^13,14^ The selection of classical ML versus deep learning methods may be influenced by the structure and size of the data. For example, deep learning models are better suited to handle high-throughput, multimodal data streams, such as raw sensor time series,^15,16^ but typically require larger datasets than classical statistical or ML methods (in terms of both features and sample size). Finally, clinical wearable data may be collected over several seconds to minutes, and longer in passive monitoring settings; thus, appropriate understanding of large dataset processing is important. Collectively, both data type, size, and model selection are key components of a comprehensive wearable sensor data analysis pipeline and should be considered when evaluating clinical research pipelines with large heterogenous datasets.

In this work, we demonstrate that assessment of vertical GRF (vGRF) using state-of-the-art, research-grade force plates can be achieved with less expensive, more accessible, biosensor insoles that allow remote data collection. We assessed vGRF data from three datasets collected with the Moticon ReGo system (referred to here as a digital insole) in subjects with knee arthropathy and control subjects to establish criterion validity of the digital insole, as compared to the force plate clinical standard. Using force plate measurements, we created an ML framework for detection of knee arthropathy status. We then validated this model using independent datasets collected with the digital insole. Next, we analyzed both derived gait characteristics and raw sensor time series data from the digital insole. Through an integrated analysis of these various data sources we identified disease signatures for knee arthropathy and detected individual-specific gait patterns. These results provide a framework for meaningful clinical interpretation of digital biomarker data and support their potential broader role for future application as clinical trial endpoints.

## Results

### Platform-agnostic visualization of knee arthropathy signatures from vGRF data

vGRF is the major and clinically relevant component of the ground reaction forces generated from walking and can be measured using force plates or digital insoles. We obtained vGRF data from the three studies: The GaitRec Force Plate study of subjects with knee injuries (*N* = 625) and healthy controls (*N* = 211) (**Supplementary Fig. 1**) and two digital insole [Moticon ReGo AG] studies in healthy controls (*N* = 22) and subjects with knee OA (*N* = 40), respectively (**Fig. 1**). We plotted raw and normalized vGRF values recorded by force plate and digital insole devices and observed by visual inspection that the means of vGRF curves within each disease category are similar across platforms (**Fig. 2a**). We observed distinct patterns between control subjects and those with knee arthropathy, when comparing the normalized vGRF values averaged by group (**Fig. 2a**) or per individual (**Fig. 2b**). Specifically, individuals with knee arthropathy from all evaluated datasets displayed a qualitatively different gait signature than healthy controls, with a “ flatter” vGRF curve shape during the middle of stance phase (we use the term ‘arthropathy’ to encompass both knee injury and OA). Using a dimension reduction approach Uniform Manifold Approximation and Projection (UMAP) with normalized vGRF data from each subject in two dimensions, subjects at a population level separated out by arthropathy status (knee arthropathies vs controls) rather than by measurement platform (**Fig. 2c**). Thus, despite performing analysis on data collected from different devices at different sites, we could discern disease-relevant patterns in the vGRF data and show that the digital insole data recapitulated the force plate data.

**Fig. 1.**
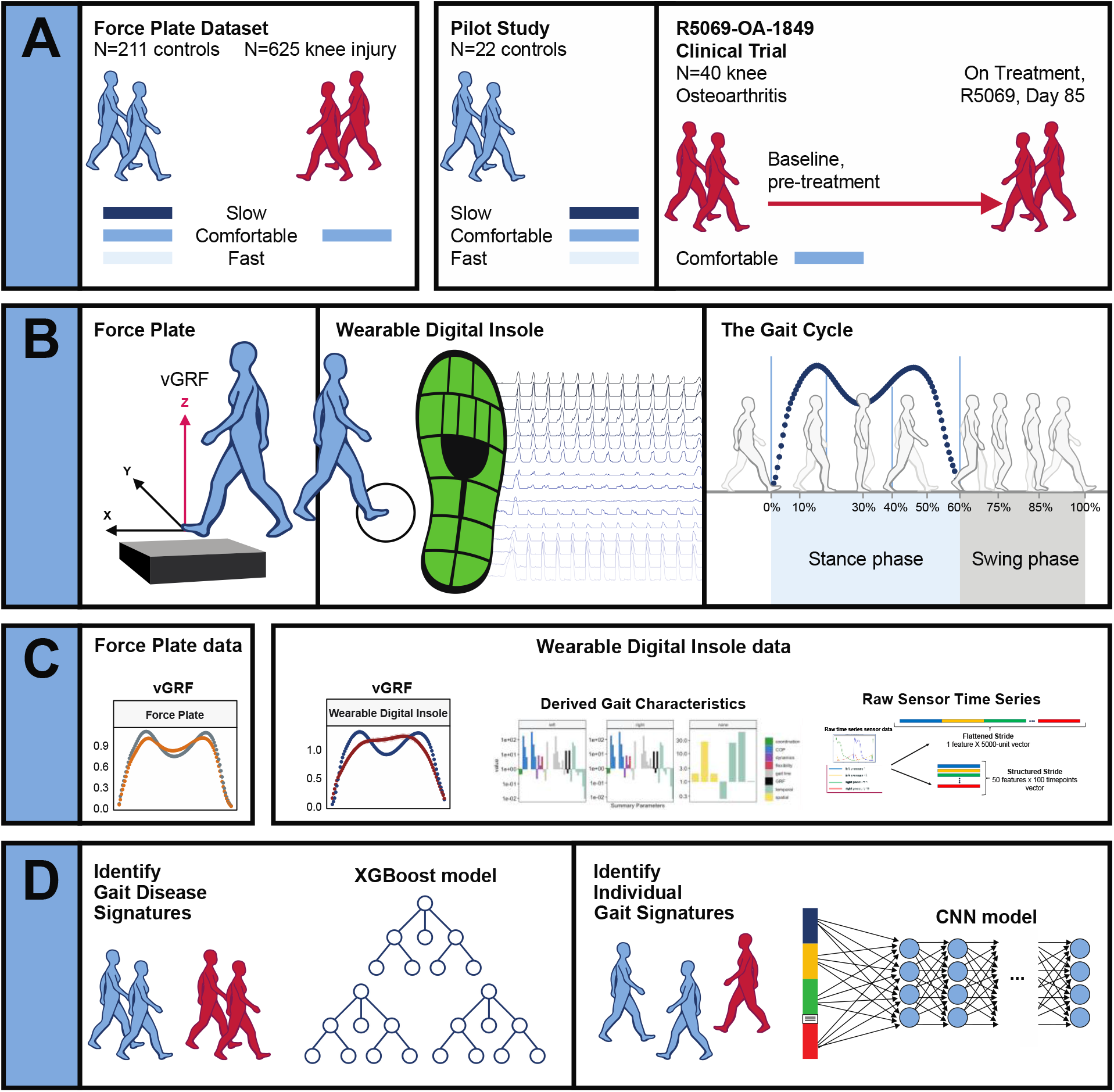
Overview of data sources and study participants, device types, data types, and clinical research questions. **a**, Three datasets were used for analyses. The GaitRec force plate dataset (force plate data) contains *N* = 211 healthy controls, who walked at three different walking speeds (slow, comfortable, and fast), and *N* = 625 knee injury subjects, who walked at a comfortable walking speed.25 The second dataset is from a digital insole pilot study, where *N* = 22 healthy controls walked at three different walking speeds (slow, comfortable, and fast). The third dataset is from a digital insole sub-study from a longitudinal clinical trial in knee osteoarthritis (OA), where *N* = 40 knee OA subjects performed a 3-minute walk test (3MWT) at a comfortable walking speed at baseline (pre-treatment) and at day 85 (on treatment). **b**, Both force plates and digital insoles produce data collected during stance and swing phases of a person’s gait cycle **c**, Types of data produced by these devices include vertical ground reaction force (vGRF), derived gait characteristics, and raw sensor time series. **d**, Clinical research questions addressed in this work include the derivation of gait disease signatures of knee OA and investigation of the individuality and consistency of gait patterns. Two analytical methods were used to evaluate these data. XGBoost, a gradient boosting classifier, was used to analyze vGRF, derived gait characteristics, and raw sensor time series flattened stride data. A one-dimensional convolutional neural network (CNN) was used to analyze structured stride raw sensor time series data.

**Fig. 2.**
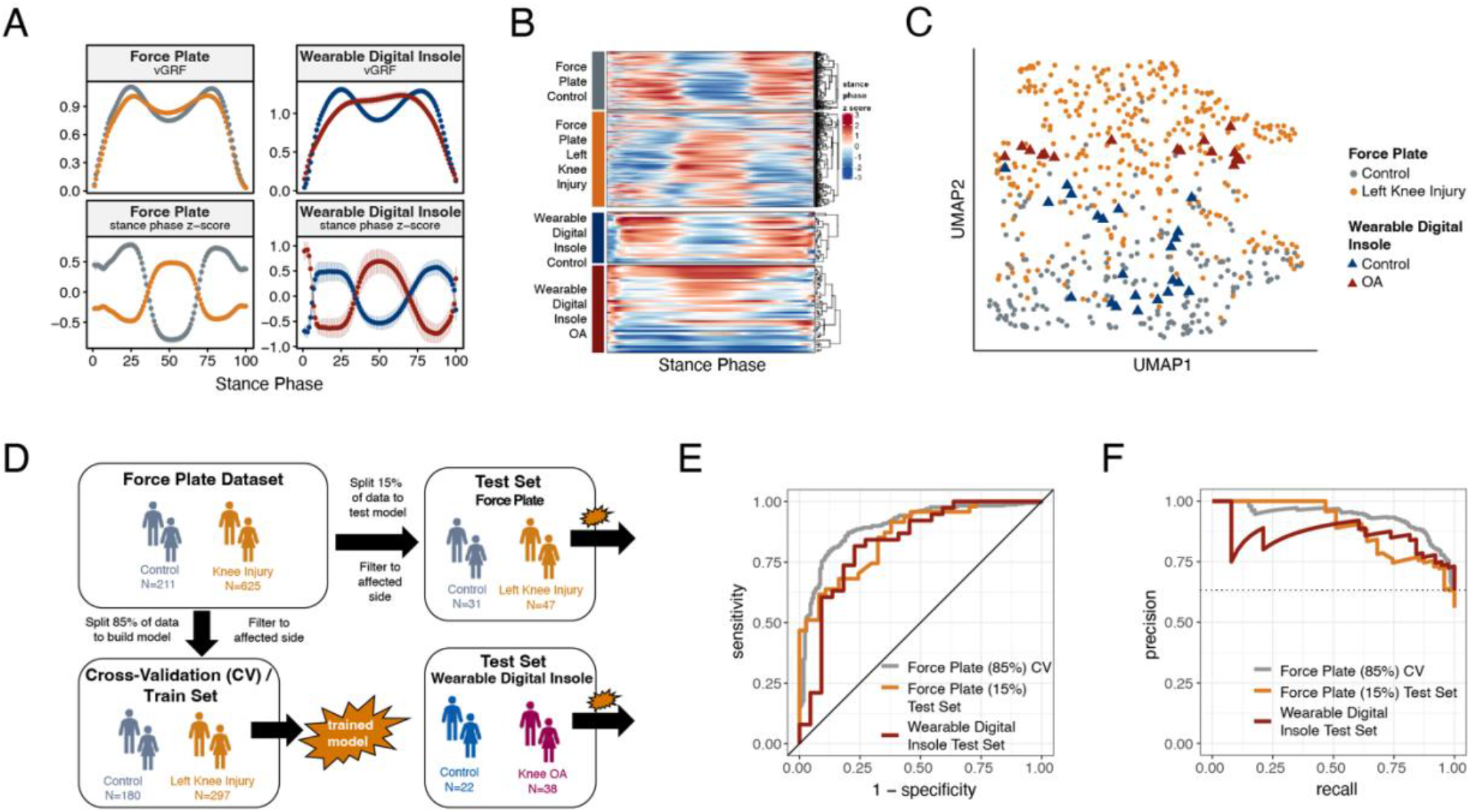
ML model trained on knee injury subjects walking on force plates accurately classifies OA patients wearing digital insoles. **a**, vGRF curves derived from force plate and digital insole data for healthy controls, and knee injury and knee osteoarthritis (OA) patients respectively. Left foot data are shown as mean of values (top panels) and mean of normalized z-scores (bottom panels) at each percent stance phase within each device and health status. Groups are color-coded as in B and C. **b**, Vertical ground reaction force (vGRF) curves for an individual’s left foot shown as heatmap rows, after data was z-transformed at each percent stance phase (as in A). Rows are hierarchically clustered within each group of subjects. **c**, Uniform Manifold Approximation and Projection (UMAP) dimensionality reduction of the z-transformed left foot vGRF data. Each point represents a subject, and points are colored by phenotype, and shaped by device. **d**, Schematic of machine learning model building of training/validation and testing sets. Two XGBoost models were created, one for left knee injury (depicted) and one for right knee injury. The full force plate vGRF dataset with both controls (comfortable walking speed) and left or right knee injury subjects (comfortable walking speed, excluding subjects with knee injury on both joints) were split 85% into training/validation datasets, and 15% into a hold-out testing set. One model predicts control versus knee injury subjects using left foot data (of left knee injury subjects and all controls), and the other predicts using right foot data (of right knee injury subjects and all controls). These models were then applied on a separate, independent testing set of digital insoles vGRF data with *N* = 22 control subjects and *N* = 38 patients with knee OA. **e**, Receiver operating characteristic curve for XGBoost classification of force plate (85%) cross-validation (CV, training/validation) set, force plate (15%) hold-out test set, and the digital insole test set. **f**, Precision-recall curve for XGBoost classification of the same groups in e.

To investigate how the variation in the vGRF data may be partially explained by the clinical and demographic characteristics of the participants, we fit a series of linear models to each point along the vGRF curve, with arthropathy state (knee arthropathy or control), age, sex (male or female), and body weight as covariates in the model (**Supplementary Fig. 2**). For each linear model, representing the sum of squares for each category compared to the total sum of squares as a percentage of variation explained by that component, we observed that the disease state is the major contributor to the vGRF signal for most of the vGRF curve, with age, sex, and body weight explaining a smaller proportion of the variance. We conclude that the dominant factor likely contributing to variation among participants to these signals is arthropathy state.

### ML models trained on vertical ground reaction force plate data to classify control versus knee injury across different platforms

We next looked to quantify how well force plate data can identify disease using gait signatures, and to understand if a wearable insole could detect similar characterizations. To differentiate controls versus knee arthropathies using vGRF data, we divided the complete vGRF force plate dataset into a training/validation set (85%) and a test set (15%) and trained an XGBoost model (gradient boosting ML model) to predict these classes (**Fig. 2d**). The model indicated strong predictive power to classify control versus knee injury subjects using force plate data, when evaluated using five-fold cross validation of the training/validation dataset, a standard method of assessing model performance (left knee: area under the receiver operating characteristics curve [auROC] = 0.90, area under the precision-recall curve [auPR] = 0.93; right knee: auROC = 0.94, auPR = 0.96). The predictive power was also strong when assessed on the hold-out test dataset, which was not used for training of the XGBoost model (left knee: auROC = 0.86, auPR = 0.90; right knee: auROC = 0.86, auPR = 0.92) (**Table 1, Supplementary Fig. 3**).

**Table 1:**
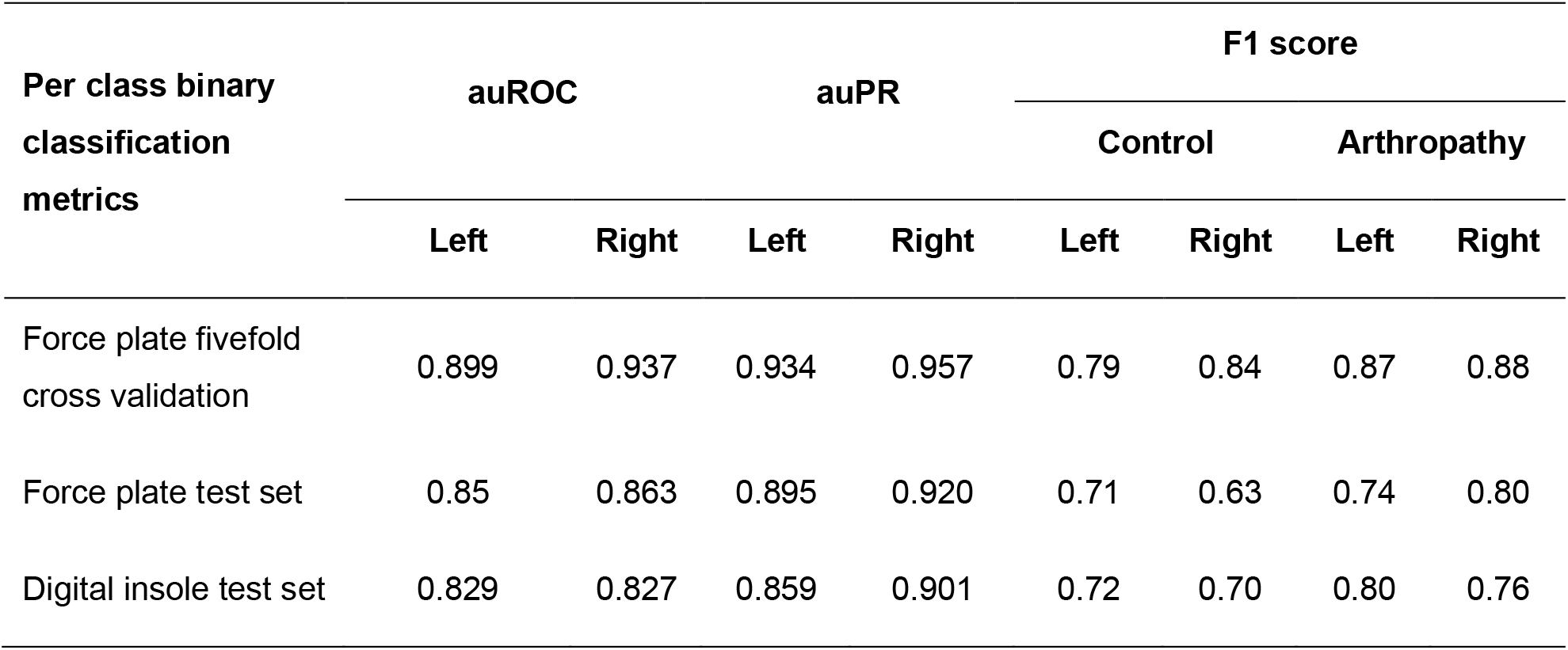
Force plate vertical ground reaction force (vGRF) control versus knee arthropathies XGBoost classification model evaluation statistics (left foot / right foot). An XGBoost model was trained on 85% of the force plate dataset vGRF data to predict control or knee arthropathies (knee injury or knee osteoarthritis) classes, with left foot vGRF data used to predict left knee arthropathies and right foot vGRF data used to predict right knee arthropathies). The model was evaluated using fivefold cross validation, a hold-out force plate test set, and a digital insole test set. Area under the receiver operating characteristics curve (auROC) and area under the precision-recall curve (auPR) statistics are reported for the three models. F1 scores for each class for each model are also reported.

| Per class binary<br>classification<br>metrics | auROC |  | auPR |  | F1 score |  |  |  |
| --- | --- | --- | --- | --- | --- | --- | --- | --- |
|  |  |  |  |  | Control |  | Arthropathy |  |
|  | Left | Right | Left | Right | Left | Right | Left | Right |
| Force plate fivefold<br>cross validation | 0.899 | 0.937 | 0.934 | 0.957 | 0.79 | 0.84 | 0.87 | 0.88 |
| Force plate test set | 0.85 | 0.863 | 0.895 | 0.920 | 0.71 | 0.63 | 0.74 | 0.80 |
| Digital insole test set | 0.829 | 0.827 | 0.859 | 0.901 | 0.72 | 0.70 | 0.80 | 0.76 |

To further assess generalizability of the model and understand if a digital insole could measure vGRF similarly to a force plate, we applied the model trained using vGRF force plate data to the separate, independent dataset derived from digital insole studies on individuals with knee OA and on healthy controls. An important assumption of this analysis was that knee OA and knee injury are represented similarly by vGRF curves as was shown earlier (**Fig. 1a-c**). We found that for the digital insole, an XGBoost model trained only on force plate data performed nearly as well on the digital insole data (left knee: auROC = 0.83, auPR = 0.86; right knee: auROC = 0.83, auPR = 0.90) as it did on the hold-out force plate test data (**Fig. 2d,e; Table 1, Supplementary Fig. 3**).

### Performance of derived gait characteristics of the digital insole versus vGRF and raw sensor time series for disease classification

A potential benefit of a digital insole relative to a force plate is that many more variables can be derived, thus enabling a more comprehensive assessment of gait. For instance, in addition to the vGRF curves, derived gait characteristics and raw sensor time series data can be obtained using the 50 sensors across both insoles (**Fig. 3a**).

**Fig. 3.**
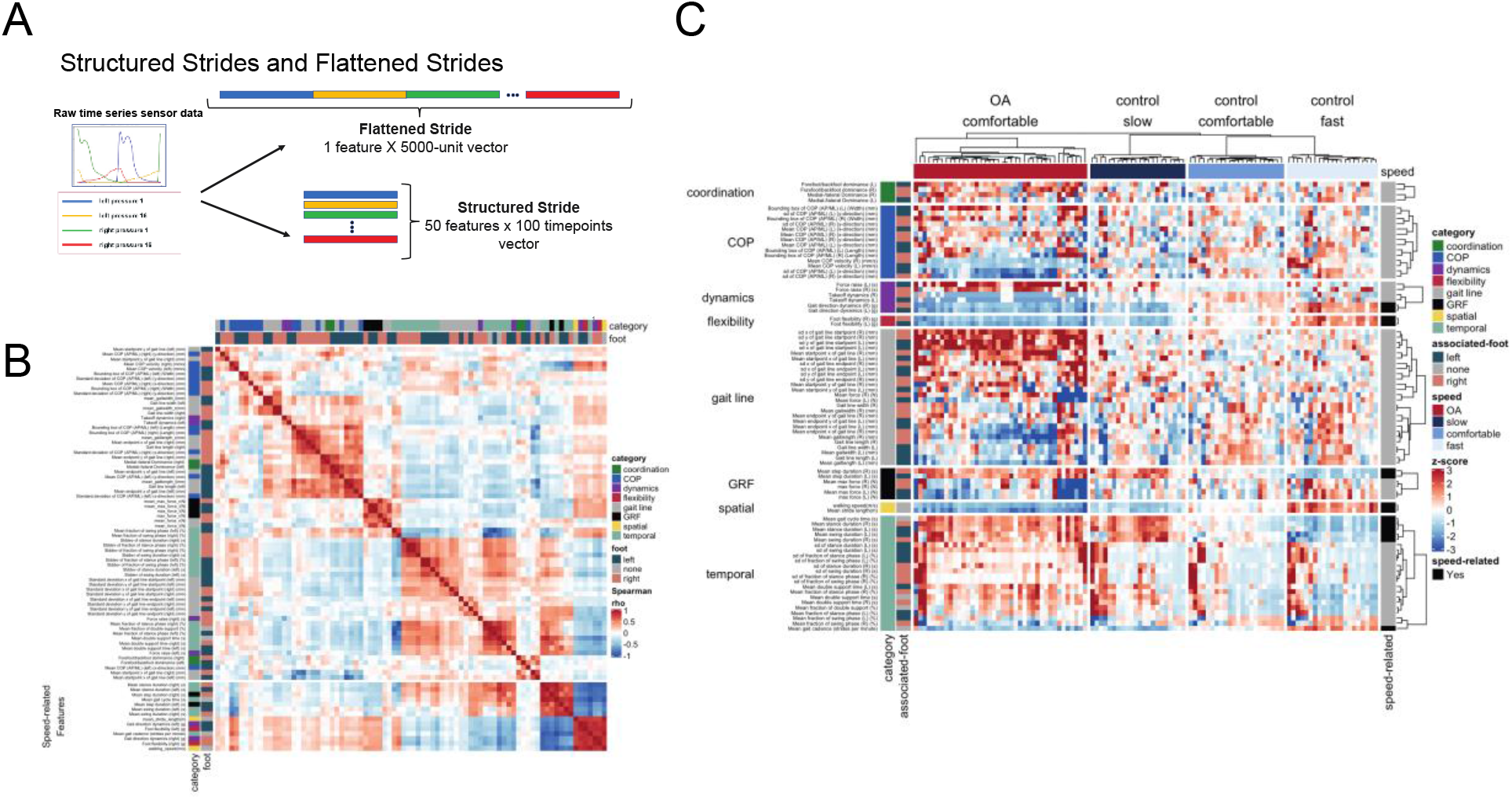
Derived gait characteristics from a digital insole measured across all subjects from the pilot study and knee osteoarthritis patients in the clinical trial. **a**, Schematic of raw sensor time series data from a digital insole. Data can be processed from the device in three ways: (1) vertical ground reaction forces (Fig. 1); (2) derived gait characteristics on force, spatio-temporal, and center of pressure aspects; and (3) raw sensor time series data from the 50 sensors embedded across both insoles. Each segmented stride of raw sensor time series data can be analyzed as is (structured strides) or collapsed (flattened strides). **b**, The derived gait characteristics (parameters) of the digital insole from all individuals in the pilot study were correlated against each other at the comfortable walking speed. Spearman correlation coefficients were computed and shown in a correlation matrix ranging from –1 (perfect anti-correlation) to +1 (perfectly correlation). Each parameter has a Spearman correlation coefficient of +1 with itself (red diagonal). The parameter, the foot from which it was generated, and its category are labeled on the left of the correlation matrix. **c**, Heatmap representation of the average of each of the 82 digital insole parameters (rows) across all walks for each patient (columns) from the pilot study. Parameter values are shown as normalized z-scores (bounded within ±3), calculated across all participants, and walking speeds. The heatmap is split by the three walking speeds (slow, normal, fast), and columns are clustered within each walking speed using hierarchical clustering with Euclidian distances. The 14 parameters strongly correlated with walking speed are indicated on the right of the heatmap.

Time series data measures different aspects of a stride, including force, angular velocity, and orientation of the foot relative to gravity. The raw sensor time series data can be evaluated as either structured strides, in which each stride is represented such that each sensor’s values are measured over time, or converted into flattened strides, in which all timepoints and sensors are concatenated into one representation (**Fig. 3a**) providing a linear representation of a persons’ stride.

To investigate how derived gait characteristics relate to each other, we clustered the values and their correlations across different walking speeds and disease status (OA vs control). Correlations within and between categories of parameters revealed that similar groups of parameters clustered together (**Fig. 3b**), including 14 derived gait characteristics correlating with walking speed. Using a heatmap, where derived gait characteristics were normalized across both control and knee OA populations, we observed distinct patterns between derived gait characteristics at different walking speeds and disease status (**Fig. 3c**). To further explore the relationship with walking speed, we performed a principal component analysis (PCA) dimensionality reduction on each data type. This analysis demonstrated that knee OA arthropathy state can be observed on a continuum related to walking speed. Compared to control subjects, participants with knee OA are walking more slowly as apparent across all data types, including vGRF (**Fig. 4a**), derived gait characteristics (**Fig. 4b**), and raw sensor time series data (**Fig. 4c**).

**Fig. 4.**
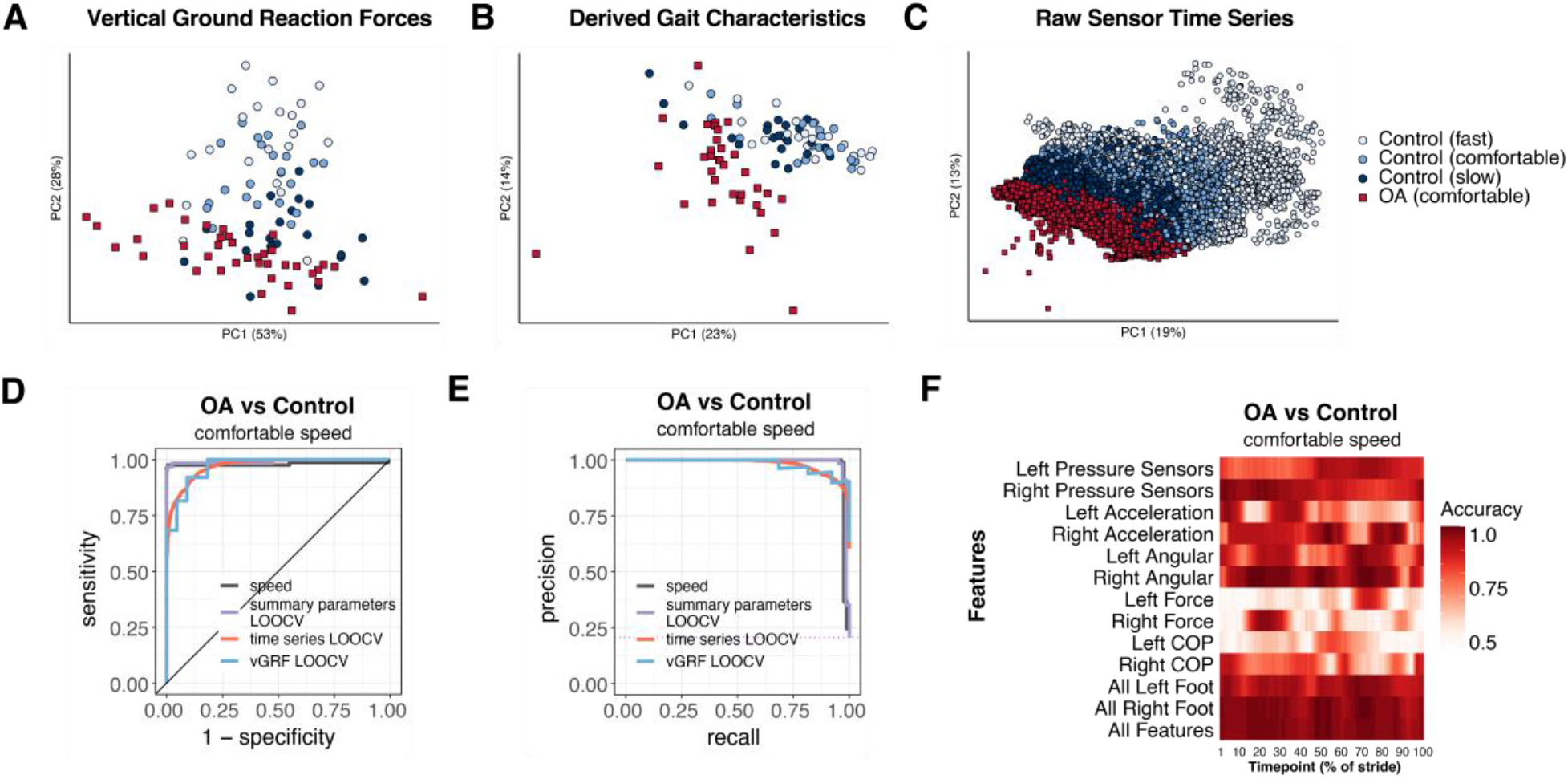
Different methods to analyze control subject versus knee OA patient data from a digital insole enable refined classification of disease signatures. **a**, Principal component analysis (PCA) dimensionality reduction of vertical ground reaction force (vGRF) data from all walks of pilot study subjects and baseline walks of knee osteoarthritis (OA) clinical trial patients. Each dot represents data from a single subject at a given walking speed. **b**, PCA dimensionality reduction of derived gait characteristic data from the digital insole, without the 14 speed-correlated derived gait characteristics. **c**, PCA dimensionality reduction of raw sensor time series of each stride from all walks. Each dot represents data from a single stride and repeat strides from the same participant are shown. **d**, Receiver operating characteristic curves for knee OA versus control (both at comfortable walking speed) prediction using only walking speed (speed), derived gait characteristics (excluding 14 speed-correlated features), raw sensor time series, and vGRF. Classification metrics were derived using leave-one-out cross-validation (LOOCV). The single derived gait characteristic speed separates out digital insole knee OA patients versus control subjects. **e**, Precision-recall curves of the same comparisons in d. **f**, Classification accuracy using raw sensor time series data from control subjects versus knee OA patients using subsets or all 50 sensors at each timepoint of the stride (0–100% of the stride). Timepoints start with the stance phase of the right foot and swing phase of the left foot, and end with the swing phase of the right foot and the stance phase of the left foot. Classification accuracy of 1.0 indicates perfect knee OA versus control classification, using data from that timepoint.

We revisited the question of whether gait data can be used to identify arthropathy status relative to control subjects, for each of the three types of data collected by the digital insole: vGRF, summary parameters, and time series data. Note, this analysis aims to classify whether a subject has knee arthropathy, rather than to determine the severity of arthropathy, which this study was not designed to assess. XGBoost models were trained and assessed using leave-one-out cross-validation (LOOCV) on vGRF, where models are evaluated by iteratively leaving one subject out, building a model, and evaluating where that subject would be classified compared to the true result (see Methods). This was performed for derived gait characteristics, and raw sensor time series (flattened strides) data independently (**Fig. 4d,e**). Additionally, we used walking speed as a single variable predictor of knee OA (both healthy subjects and OA patients had been asked to walk at a self-paced comfortable walking speed). We found that speed alone was able to discriminate between knee OA subjects and healthy controls (auROC = 0.981, auPR = 0.983). vGRF LOOCV with the digital insole also demonstrated high predictive power (auROC = 0.970, auPR = 0.982). For derived gait characteristics, we wanted to understand whether aspects of gait other than walking speed could be used to correctly classify whether a subject had knee OA relative to a control as this would suggest potential for broader applicability if disease specific features in addition to changes in speed could be detected with digital insoles. Using derived gait characteristics that excluded the 14 characteristics correlated to walking speed, we found even better classification accuracy (auROC = 1.000, auPR = 0.999). The most important discriminating parameters included Takeoff dynamics, max force (N), Mean COP velocity (mm/s), and gait line associated parameters (sd x and sd y of gait line start point (mm) (**Supplementary Fig. 7**). Flattened strides from raw sensor data, which is also independent of walking speed as each stride was interpolated to a consistent 100 timepoints, also was predictive but with slightly less accuracy (auROC = 0.972, auPR = 0.981). (**Supplementary Fig. 4**)

Finally, we sought to evaluate the contribution of each sensor at each timepoint along individual segmented strides to the disease classification accuracy. We trained additional XGBoost models on subsets of sensor type at each timepoint (**Fig. 4f**) and found that classification accuracy for control versus knee OA depends on the type of sensor, the timepoint along a stride, and the foot (left vs right). Measurements of pressure, force, and angular rate from a foot had greater mean classification accuracy during the stance phase of that foot.

### Deriving individual gait signatures using convolutional neural net latent representations of raw sensor time series data or derived gait characteristics

To determine the extent to which walking patterns are specific to an individual, subjects were split 50:50 into training and testing sets and stratified by disease status (**Fig. 5a**). We then trained a one-dimensional convolutional neural net (CNN) on structured strides of training set individuals, and subsequently applied the CNN model on structured strides of testing set individuals. For each stride, we extracted the 60 features in the last connected (penultimate) layer of the CNN (**Fig. 5b**). This layer directly precedes the final output of the CNN model predicting the individual from which the stride came, and thus these 60 features constitute a gait “ fingerprint” learned by the CNN model. These features were learned by the CNN to distinguish individuals, and thus these patterns (captured in the latent features) can subsequently be used for classification of new subjects, previously unseen by the model.

**Fig. 5.**
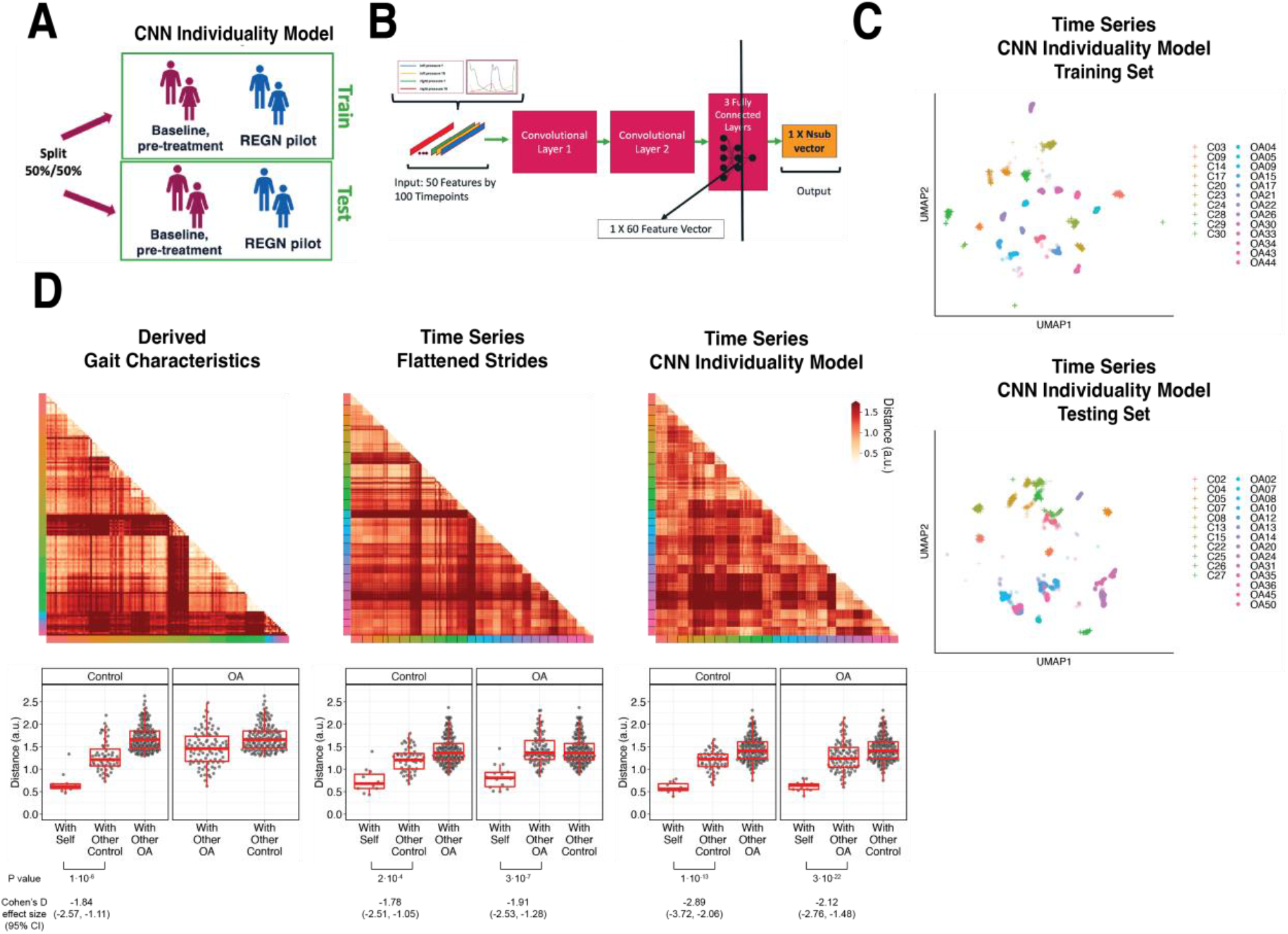
Latent CNN representation of raw sensor time series data from digital insoles: identifying subject-specific patterns of human gait. **a**. Pilot study subjects and knee osteoarthritis (OA) clinical trial patients were split 50:50 into training and testing sets, stratified by disease status, for the first convolutional neural network (CNN) model investigating the individuality of gait patterns. **b**, A CNN was trained on segmented structured strides from the digital insole in the training set, to predict from which subject the stride came. The activation of the last fully connected layer in the CNN consists of 60 features and represents the model’s latent representation of gait. **c**, Uniform Manifold Approximation and Projection (UMAP) clustering of these 60 latent features for each stride captures the individuality of participants in both the training and testing sets. Each dot represents a single stride, colors represent each participant, and shapes represent participants’ health status (C = control). Intra- and inter-subject clustering and separation is greater in the training set, as expected, and is present in the testing set as well. **d**, Distances (in arbitrary units) between each pair of walks (for derived gait parameters) or strides (for time series) from the testing set shown as heatmaps for each of the three methods (top panels). Subject of the walk/stride are color identified along the edge. Boxplot of mean distance of each walk/stride with other walk/strides from the same individual, and with walk/strides from other individuals separated by disease class (bottom panels). Distances are faceted by the disease class of the individual. A good representation has low distance for “ with self”, and high distance for “ with other” classes.

To visualize the latent representation of strides from the CNN, a UMAP clustering of these latent features from each stride indicated that this representation captured the individual identity of participants (**Fig. 5c**), as strides from the same individuals clustered together in both the training and testing sets.

We sought to quantify the individuality of each of the three representations of gait: derived gait characteristics from each walk, raw (flattened) time series of each stride, and CNN latent features of each stride. To do so, we calculated the distance between all pairs of test set walks/strides in such representation against each other (see Materials and Methods). These distances are displayed in both heatmaps (**Fig. 5d**, top) and in boxplots (**Fig. 5d**, bottom). An ideal representation to quantitate individuality would have low distances between walks/strides from the same person, and high distances for between walks/strides from different people (**Supplementary Fig. 5**).

Subjects within their class (control or knee OA) displayed similar gaits therefore, we separated out comparisons of subjects to their same class versus comparisons to the other class (with other control, with other OA). All three representations had significant differences in distances when comparing strides from the same subjects versus strides from different subjects from the same class (*P* ≤ 0.0002, two-sided t-test). The CNN latent representation was best at minimizing distances of strides from the same subjects while maximizing the distances of strides from different subjects from the same class, as measured by Cohen’s d effect size, suggesting that the CNN latent representations best capture gait individuality, and as such was used for follow-up analysis.

### Evaluation of individual gait signatures after training on raw sensor time series from multiple days

A second CNN model was trained on combined data from both timepoints in the R5069-OA-1849 clinical trial, where input data was labeled only by participant and not by timepoint. We call the first model trained only on day 1 data an “ individuality” model, and the second model trained on both day 1 and day 85 a “ consistency” model (**Fig. 6a**).

**Fig. 6.**
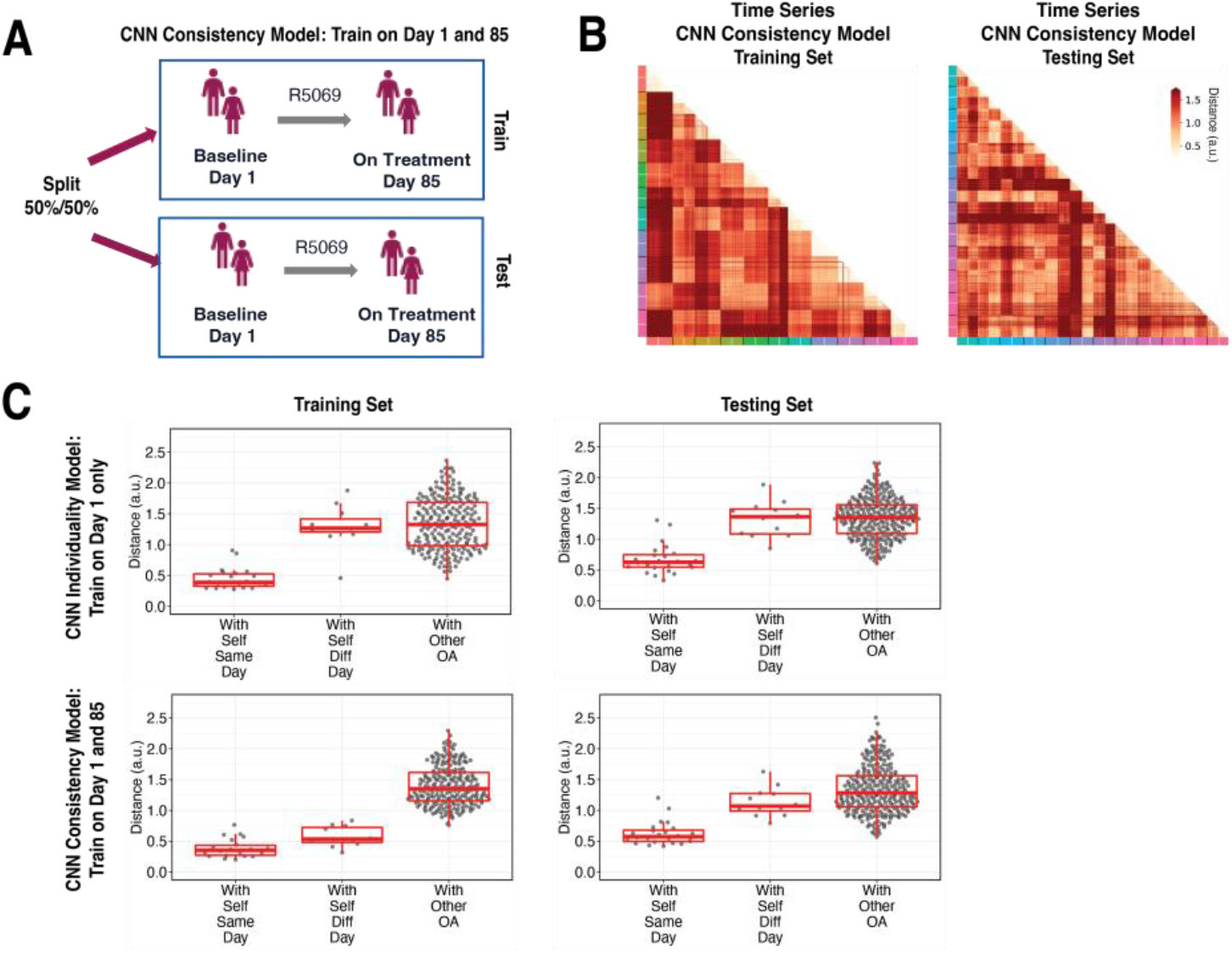
Training across multiple days increases consistency of CNN model latent representation. **a**, Knee osteoarthritis (OA) clinical trial participants were split 50:50 into training and testing sets containing both day 1 (baseline) and day 85 (on treatment) data, for the second convolutional neural network (CNN) model investigating the consistency of gait patterns. **b**, Distances (in arbitrary units) between pairs of strides in the latent representation from the consistency CNN model in the training and testing sets, shown as heatmaps. Strides from the same person are arranged next to each other, with strides from day 1 listed first then strides from day 85. Color along the edge indicates each person. **c**, Boxplots of mean distance of each stride with other strides from the same person on the same day, from the same person on different days, and from other people, for both the individuality model (Fig. 5) and consistency model (a–b). Distances are shown using the different models in both the training and testing sets.

We tested both models on day 1 and day 85 by evaluating the distance of the CNN penultimate layer of all strides with each other. **Fig. 6b** plots the distance for the consistency model on both the training (left) and testing set (right) participants. For each participant, day 1 and day 85 strides are arranged next to each other, and the cross-day distance within each participant are shown in squares closest to the diagonal.

**Fig. 6c** shows stride distances for within subject from the same day, within subject from different days, and within other subjects. Within the training set, the consistency model, which was trained on both day 1 and day 85 strides, produced lower distances than the individuality model, which was trained on only day 1 strides, in comparing strides within self from different days (*P* = 3·10^−5^, two-sided paired t-test) (**Supplementary Fig. 6**). Importantly, within the testing set, the consistency model produced lower distances than the individuality model in comparing strides within self from different days (*P* = 0.033, two-sided paired t-test), suggesting that training across multiple days improves the ability of the CNN consistency model to identify features that remain consistent across multiple visits. The results also show additional capacity for model improvement with respect to consistency of gait, as in the consistency model, the distances of the data to oneself were lower amongst strides from the same day versus different day in both training and testing participants (*P* = 4·10^−3^ and *P* = 4·10^−6^, respectively, two-sided t-test).

## Discussion

Digital health technologies, which include wearable devices, generate enormous amounts of data, even if worn for short periods of time. While in theory these data are no different than other types of biomarker data when generated in clinical research settings, its structure, dimensions, size, and interpretability present unique challenges for clinical research. In this work, we investigated how data derived from a digital insole can answer questions of clinical interest (with the goal of yielding endpoints), including the detection of disease specific patterns relative to controls, and the identification of subject-specific gait patterns.

To address the question of how to estimate disease specific patterns, we tested whether vGRF data allowed for detection of knee arthropathy relative to control subjects with any device. We built an ML model using force plate vGRF data and tested the model on an independent force plate test dataset. We then extended the analysis to the data from the Moticon digital insole. We observed consistent disease versus control differences in vGRF curves with both technologies, such that these models were able to distinguish knee arthropathy subjects from their respective control groups. The predictive performance of the ML model on data collected from a different device at different physical locations and experimental conditions suggests that the model was robust and generalizable (not overfit). While our analysis may be confounded by variables like age, our results imply that disease status is the major signal we observe. Our results suggest that vGRF data likely reflect true differences between control and knee arthropathy subjects, suggesting that digital insoles may be used to screen for knee arthropathy and potentially other diseases that impact gait.

We next sought to identify how different types of digital biomarkers beyond vGRF (where the digital insole attempts to replicate exactly what is measured on a force plate) can be utilized. While force plates have notable advantages in terms of accuracy, the ease and generalizability of using wearable devices in larger populations or trial settings where specialized gait labs are not accessible can enable more widespread implementation of gait analysis. To provide additional insight into the advantages of utilizing wearable devices, we investigated alternative data types generated by wearable devices.

Evaluation of derived gait characteristics from the digital insole highlighted walking speed as an important determinant of knee OA classification, which is expected;^17^ however, when derived gait characteristics highly correlated with speed were removed, the model still successfully detected knee OA subjects. Despite walking speed being clinically relevant^18-21^ in studies of overall health status, it is unlikely to be able to distinguish between various musculoskeletal and neurological diseases and a speed-independent approach could enable more informative evaluation of gait disease signatures. This highlights that in addition to alterations in spatiotemporal parameters, there are additional features of gait in knee OA subjects that differentiate them from controls.

Knee OA contributes to altered gait since individuals try to avoid knee pain and cartilage contact stress. A recent meta-analysis showed that biomechanical alterations associated with knee OA during level walking were significantly increased lateral trunk lean toward the ipsilateral limb along with non-significantly increased trunk/pelvic flexion and resultant significant alterations in external hip adduction moments.^22^ However, the body of evidence for these outcomes was ranked very low. Issues included a risk of bias, imprecision, and heterogeneity due to patients’ characteristics and methodological differences in gait analyses. Our work confirms that such speed independent characteristics do indeed play an important role in identifying gait of patients with knee arthropathy from controls in an objective manner.

Distinguishing between healthy and disease subjects, where effect sizes are expected to be large, may have important clinical implications. However, it remains to be seen whether wearable devices can successfully detect disease severity where effect sizes are smaller. Our study was not designed to investigate smaller effect size differences, so future studies are needed to evaluate this further. If future studies show this is feasible, this data would be important to determine disease progression or evaluate the impact of therapeutic interventions, and could have utility in clinical practice, as well as serve as an endpoint tool for clinical trials. The objectivity of the data also makes these suitable for endpoint tools. For example, derived gait characteristics could represent reliable registrational endpoints in clinical research, given that they describe objective aspects of gait with clinical relevance (e.g., total distance walked in meters, or maximum force applied during a 3-minute walk in Newtons).

Finally, evaluation of raw time series data from digital insoles demonstrated that data from a single stride could identify individual subjects. However, the interpretation of time series data remains challenging, particularly when analyzed with deep learning methods. In current clinical trial settings, derived gait characteristics may be a simpler approach. Nevertheless, our finding that raw time series data contain subject-specific latent features suggests that potentially useful gait features, beyond the disease signatures studied here, may exist in this data.

Collecting additional timepoints from individuals may permit the model to learn more consistent subject-specific gait patterns. Individual subject gait patterns have been reported previously,^13,23,24^ and understanding their quantification may be useful in clinical development for precision medicine applications. Subject-level gait patterns and the ability to identify unique signatures of an individual’s gait may enable improved monitoring of treatment responses on a per-subject as well as on a population-wide level. Training datasets with participant data from multiple visits improves the ability of the model to detect gait features that remain consistent, or that change, with time. Collection and training on additional timepoints beyond the two days in our study may result in models that better learn gait features to consistently identify an individual across time.

This work highlights the potential utility of leveraging devices for clinical research though originally developed for other purposes, such as the Moticon digital insole, which was developed for athletic sports training. However, the data such devices generate require clear hypothesis-driven validation to detect relevant signals just like research-grade instrumentation. Holistically, by showing that vGRF data from a digital insole replicate the standard clinical data generated from force plates, we demonstrated criterion validity of vGRF data from digital insoles, meaning that digital insoles can to some degree replicate a clinical standard (the criterion). We further demonstrated the external validity within the digital insole study of the disease gait signature across both methodologies (force plate and digital insole) using an ML approach, with a training set built entirely on force plate data and evaluated on both force plate and digital insole data collected elsewhere. Analytical strategies that maximize both clinical understanding and generalizability to other studies are of course standard for biomedical research.

However, we go beyond this step, and further attempt an analytical approach to maximize construct validity—how close a digital biomarker reads out the “ construct” it is intended to measure—even at the expense of face validity (the degree to which a measure is intuitively interpretable), for a particular clinical question. Here, the fact that raw sensor data lacks face value interpretability but improves upon our ability to ascertain subject-specific gait patterns may inform us that these digital biomarker data contain disease or subject specific information that could be leveraged in alternative clinical circumstances.

Limitations of this analysis include the small sample size in the pilot study of healthy controls. Despite this, we were able to show favorable classification performance for these control subjects relative to their force plate counterparts. While this study only looked at knee arthropathies, future work will be focused on a broader set of gait-affecting diseases. While only two repeat timepoints for subject-level classification were available in the knee OA group, we demonstrated a modeling approach to reliably compute an individual’s gait consistency. In the future, this would ideally be performed on data collected at more than two timepoints. In addition, derived gait characteristic level data were not available for the force plate dataset to compare with the digital insole generated data.

In summary, this work outlines a framework for an integrated analysis of digital insole data to answer clinical and research questions relevant to digital endpoint development. To identify disease signatures, we built an ML model using only data from force plates, the clinical standard, and analyzing data from a digital insole, we showed comparable disease classification. This platform-agnostic analysis demonstrates that ML approaches can help support and validate digital biomarkers and yield digital endpoints of clinical utility. In addition, the finding that our models can identify individual gait patterns suggests that data generated from a digital insole may have unforeseen future applications such as the potential to detect changes in individual gait patterns, which may provide better understanding of the impact of a therapeutic intervention for that individual. Ultimately, this technology may provide value in the health care delivery setting, aiding in accurate diagnosis or longitudinal monitoring of disease progression or of response to treatment.

## Data Availability

Anonymized data and computer code to reproduce all figures will be made available as a supplementary file to this manuscript when published. The GaitRec dataset is available online here: https://www.nature.com/articles/s41597-020-0481-z

## Acknowledgements

The authors would like to give a special thanks to the R5069-OA-1849 clinical trial participants who volunteered for the Moticon digital insole sub-study. The authors would like to thank Mark Waterlow, Scientific Director at Prime, for writing, formatting, and editorial assistance. We would also like to thank Qing Zhou in Regeneron Medical Affairs for editorial and project management support. We thank Daniel Choka from the Regeneron Creative Services Department for assistance with creating the figures. The authors would like to give a special thanks to Dr.-Ing. Robert Vilzmann, CTO, Moticon ReGo AG for helpful discussions throughout the development of this work.

## Funding

This study was funded by Regeneron Pharmaceuticals, Inc.

## Author contributions

Conceptualization: MFW, AZL, KG, BL, RA, SCH, OH

Data collection: XW, RA

Performed Analysis: MFW, AZL, KG, BL, ML, IS, MT, DD, FW

Reviewed Analysis: AB, WKL, GH, MA, SCH

Project administration: SSK, YP, JDH, CHD

Project supervision: XW, BK

Writing – original draft: MFW, AZL, KG, SSK, OL, AA

Writing – review & editing: all authors

## Competing interests

There are pending patents related to the work presented in this manuscript. MFW, AZL, KG, SSK, XW, JI, BK, IS, DD, AB, FW, WKL, JU, GH, YP, MA, JDH, CHD, OL, AA, RA, SCH, OH are currently or have been employees and/or are shareholders of Regeneron Pharmaceuticals, Inc.

## Code and data availability

Anonymized data and computer code to reproduce all figures will be made available as a supplementary file to this manuscript. The GaitRec dataset is available online here: https://www.nature.com/articles/s41597-020-0481-z

## Materials and methods

### Study design

The objectives were to characterize data from a wearable insole device (Moticon), demonstrate their utility relative to a clinical standard, and to investigate optimal analytical methods and data types for the analysis relevant to clinical questions of interest. Three datasets were integrated for analysis (**Fig. 1**).

The GaitRec force plate vGRF dataset contained force plate control subjects (*N* = 211) and knee injury subjects (*N* = 625).^25^

Healthy control subjects (*N* = 22) from a pilot study of a digital insole conducted between July 2019 and August 2019 were included (**Table 2**). The date of first enrollment in the pilot study is 06 July 2019 and last participant visit was 05 August 2019. Those pregnant or with a body mass index above 40 kg/cm^2^ were excluded from the study. Participants were recruited internally within the Regeneron facility located in Tarrytown, NY, USA, and were provided written informed consent prior to participation. The study was considered exempt research under the Common Rule (45 CFR Sec 46.104).

**Table 2.** Baseline characteristics and gait assessments of subjects in the digital insole pilot study and patients with knee OA in the R5069-OA-1849 clinical trial digital insole sub-study.

|  | **Pilot study (N = 22)**<br>**Cross sectional** | **Clinical trial (N = 44**<br>**enrolled in sub-study, N =**<br>**43 data collected)**<br>**Longitudinal** |
| --- | --- | --- |
| **Age, years** |  |  |
| Mean | 39 | 62.75 |
| Median | 35 | 63 years |
| Range | 19–85 | 52–77 |
| **Sex** |  |  |
| Female | 11 | 28 |
| Male | 11 | 8 |
| **BMI (kg/m<sup>2</sup>)** |  |  |
| Mean | 26.2 | 34.5 |
| Median | 26.0 | 34.8 |
| Range | 20.1 – 37.0 | 26.6 – 38.9 |
| **Arthropathy class** | N/A | K-L2-3: N=23<br>K-L4: N=13 |
| **Walk test** | Walk straight for 30 seconds | 3-minute walk test |
| **Walking speed** | 3 speeds (comfortable, fast, slow) | 1 speed (comfortable) |
| **Number of walk test performed** | ~12 times (at each speed) | 1 time at baseline and<br>1-time on-treatment<br>(day 85) |
| **Total length of gait evaluation** | 20 minutes | 3 minutes |

K-L, Kellgren-Lawrence.

As part of a clinical trial evaluating the impact of a novel pain therapeutic in moderate to severe knee OA (R5069-OA-1849; NCT03956550), a sub-study of the digital insole was performed to collect data for gait assessment in knee OA patients (results from the clinical trial will be published separately). All patients were enrolled at two study sites in the USA and Moldova and the study was conducted between June 2019 and October 2020. The date of first enrollment in the R5069-OA-1849 trial 17 June 2019, and last patient visit was 29 October 2020. The sub-study targeted to enroll approximately 13 patients per treatment group to obtain data on at least 10 patients per treatment group for a total of approximately 30 patients across the entire sub-study. Eligible participants were men and women ≥40 years of age with a clinical diagnosis of OA of the knee based on the American College of Rheumatology criteria with radiologic evidence of OA (Kellgren-Lawrence score ≥2) at the index knee joint as well as pain score of ≥4 in Western Ontario and McMaster Universities Osteoarthritis Index (WOMAC) pain sub-scale score. The WOMAC score is a self-administered questionnaire consisting of 24 items divided into three subscales, where the pain sub-score is assessed during walking, using stairs, in bed, sitting or lying, and standing upright. The study protocol received Institutional Review Board and ethics committee approvals from Moldova Medicines and Medical Device Agency and National Ethics Committee for Moldova, and the Western Institutional Review Board.

### Study procedures

In the pilot study, each participant walked straight along a hallway with a hard tile floor at three different qualitative speeds for ∼12 times at each speed (∼36 walks total). For each walking trial, participants wore the digital insole inside their own shoes and were prompted to walk at a normal or comfortable speed, walk fast as if they were in a hurry (fast speed) or walk slow as if they were at leisure (slow speed). Prior to the walking trials, each participant was instructed to practice walking around to get accustomed to the insole. Participants’ clinical and demographic information were also collected prior to walking trials.

In the R5069-OA-1849 clinical trial, a total of *N* = 44 OA patients were enrolled into a sub-study of a 259-patient clinical trial. Patients were required to bring the same pair of shoes to the study site to perform a 3-minute walk test with the digital insole. Each patient performed the task twice, once at baseline and the other 85 days later post-treatment.

### vGRF data processing

To normalize the vGRF data across devices (due to the differing sampling frequencies of force plates and Moticon) and subjects, smoothing spline functions (scipy.interpolate.interp1d) were fit to vGRF timeseries sensor data from both GaitRec force plate data and Moticon computed vGRF data. vGRF curves were bounded by 0, and 100 evenly spaced timepoints across the curve were derived for each curve (to derive a % stance phase). All vGRF curves were normalized by participants’ body weight in Newtons. Within each device, the vGRF curves were further normalized using a z-transformation within each stance phase timepoint (**Fig. 2a**).

### Digital insole raw sensor time series data processing

The digital insole collects 25 100-Hz measurements for each foot (50 measurements across both feet), comprising 16 measurements from 16 vertical plantar pressure sensors, 3 x,y,z measurements from an accelerometer, 3 x,y,z measurements from a gyroscope, 1 measurement of total force, and 2 x%,y% measurements of center-of-pressure. These raw sensor time series sensor data for both the R5069-OA-1849 clinical study and Regeneron pilot study was preprocessed with custom scripts written in Python 3.6.

For the following analysis, a “ walk” was defined as data captured by the digital insole while the subject completed the researcher’s walking task (typical duration of 180 seconds for the R5069-OA-1849 clinical study and 25 seconds for the Regeneron pilot study). A “ stride” is defined as the data captured by the digital insole between the peak pressure of the right heel (the average of digital insole right pressure 1 and 2 sensors) and the next peak pressure of the right heel. A typical stride duration is 1-2 seconds, highly dependent on individual walking speed.

Data preprocessing for each subject was performed separately. First, each walk was segmented into individual strides. Since the digital insole did not collect data in regular intervals, each stride was interpolated for each of the 50 sensors to obtain 100 timepoints along the stride. Thus, each interpolated stride consists of 50 vectors (one for each sensor), and each vector is 100 units long.

For the pilot study, walks from each subject were processed individually, treating slow, comfortable, and fast walks separately. Walks without all 50 features and walks with greater than 1% missing data were excluded. For the remaining walks, any missing data was linearly interpolated (scipy.interpolate.interp1d).

Each walk was then segmented into strides, and each stride was interpolated to 100 timepoints. To segment a walk, peaks were identified in the average timeseries of the Moticon right pressure sensors 1 and 2, located in the right heel using scipy.signal.find_peaks with parameters width = 10 and prominence = 5. The walk was segmented using the peaks, and the number of measurements in each segment was calculated. Segments had that an outlier number of samples (outliers defined as 1.5*iqr +/-q3 or q1) were excluded, such that only regularly repeating segments, or strides, were analyzed. Each of the 50 features in each stride was then linearly interpolated (scipy.interpolate.interp1d) to 100 time points. Only walks with at least 10 interpolated strides were further analyzed.

Under the assumption that an individual’s strides within a walk should be highly regular to each other, each stride’s Pearson r correlation with the means of the remaining strides was computed (stats.pearsonr), and any strides with an outlier Pearson r correlation (outliers defined as 1.5*iqr +/-q3 or q1) were excluded. This process was then repeated with the remaining strides, to obtain a list of the Pearson r coefficients of each stride with the means of the other strides. The entire walk was excluded if the mean of the Pearson r coefficients fell below 0.95. This procedure was repeated one last time, across all walks by an individual at the same walking speed (slow, comfortable fast). That is, each stride’s Pearson r correlation with the average of remaining strides in all walks at the same speed was computed. Again, assuming strides within a subject and within a given walking speed should be consistent with each other, strides with an outlier Pearson r correlation were excluded (outliers defined as 1.5*iqr +/-q3 or q1). Lastly, features dependent on body weight (i.e. pressure sensors and force sensors) were normalized by the subject’s mass.

For the R5069-OA-1849 clinical trial, data were processed similarly with the following exceptions. OA patients had digital insole data collected for only two walks, on one day 1 and one on day 85, which were processed separately. Walks with >5% missing data were excluded, and an entire walk was excluded if the average Pearson r correlation fell below a Pearson r coefficient of 0.90.

### Derived gait characteristics and identification of those speed-correlated

The digital insole derives 85 gait parameters from each walk. Of those, three are directly related to the length of the walk (walking distance and left and right center-of-pressure trace length) and were excluded from further analysis, leaving 82 derived gait characteristics.

Spearman correlations between these 82 parameters were calculated across all walking speeds (slow, comfortable, fast). The silhouette method was used to determine the optimal number of clusters with the factoextra package in R with function fviz_nbclust with 100 bootstrapped samples. Since we were interested in understanding aspects of gait other than walking speed, we correlated all parameters against walking speed, and conservatively removed 14 parameters that may be influenced by walking speed in any way (|Spearman rho| > 0.7). This allowed for an investigation into gait parameters less influenced by walking speed.

### Dimensionality reduction

UMAP method for dimensionality reduction was applied using the R UMAP package with default parameters to the z-transformed vGRF data from both the force plate and digital insole datasets to investigate batch effects.

Principal component analysis of digital insole vGRF, derived gait characteristics, and raw sensor time series was performed using the prcomp function in the R stats package. Heatmaps of Moticon parameters are displayed per individual, averaged across all individual walks. All heatmaps displayed derived gait characteristics after z-transformation by row across all subjects. All clustering on heatmaps was unsupervised, within groups.

### ML model building

XGBoost models were built using vGRF, derived gait characteristics, and raw sensor time series processed data using the sklearn and xgboost packages in Python.

The force plate dataset was randomly split into 85% training and 15% hold-out test datasets. The 85% training dataset was used for LOOCV and to construct a final trained model, which was then evaluated on the hold-out test dataset. The digital insole dataset was also used as an independent dataset for evaluating the model.

Model performance was evaluated using multiple methods. ROC and precision-recall curves were used to evaluate overall performance. Additionally, we quantitated the auROC curve, which is a standard measure of classification success and describes model performance regardless of baseline likelihood for either class. In addition, we quantitated the auPR and F1-scores, which are useful for evaluating datasets with class imbalances.

### Subject-specific gait signatures

A second question we sought to investigate is whether we could determine subject-specific gait signatures. If we could train models to identify individual subjects from their walk, or from just a single stride, this may suggest that the gait data collected has richness that can identify clinical attributes beyond knee disease. We posed this question irrespective of disease state, and rather focused on identifying the optimal method to determine an individual participant’s gait pattern.

We asked approached this question via two approaches commonly used in clinical research settings ^13,26-28^. The first is regarding the individuality of human gait patterns. We sought to understand which methodology is best suited to identify generalizable patterns of any person’s gait. The second is understanding which methodology captures features of a specific individual’s gait that have consistency with time. By analogy, a facial recognition software should identify people regardless of whether they wear hats or sunglasses. To learn that these accessories are not stable features of an individual, ML models would need to be trained on images of the same individuals with and without such accessories – that is, trained on these individuals across multiple timepoints.

Similarly, we sought to understand whether training on the same individuals across multiple timepoints improves the ability of our models to detect features that identify individuals consistently with time.

### CNN model for control versus OA classification

For the control versus OA model, model performance was determined using LOOCV. A CNN was trained using all strides from all but one participant, after which the model was evaluated on all strides of that left-out participant. Each participant was used as a left-out test participant in one model, such that for *N* participants, there were *N* different CNN models each trained on the other *N*-1 participants. Each stride was labeled as to whether it came from a control or an OA participant.

For each CNN model, strides from the *N*-1 training participants were split into an 80% training set and a 20% validation set. Each feature within each stride was scaled into 0-min to 1-max range across the 100 interpolated timepoints. The normalized data from each stride was then used as the input to the following CNN architecture: (Functions in italics from the Python [v3.9.7] PyTorch [v1.8.0.post3] package torch.nn were used with the default parameters unless otherwise noted).

- First 1D convolution layer with 50 in channels, 64 out channels, and a convoluting kernel of size 3 (*Conv1d*).
- Element-wise rectified linear activation unit (*relu* in torch.nn.functional).
- 1D max pooling with a sliding window kernel of size 2 (*MaxPool1d*).
- Dropout with 0.2 probability (*Dropout*).
- Second 1D convolution layer with 64 in channels, 128 out channels, and a convoluting kernel of size 3.
- Element-wise rectified linear activation unit.
- 1D max pooling with a sliding window kernel of size 2.
- Dropout with 0.2 probability.
- Flatten data to a linear vector of 2944 elements.
- First fully connected layer with 2944 in channels and 120 out channels (*Linear*).
- Element-wise rectified linear activation unit.
- Dropout with 0.2 probability.
- Second fully connected layer with 120 in channels and 32 out channels.
- Element-wise rectified linear activation unit.
- Dropout with 0.2 probability.
- Third fully connected layer with 32 in channels and 1 out channel.
- Logistic sigmoid function (*sigmoid* in torch).

Binary cross entropy loss (*BCELoss*) was used as the loss function, and stochastic gradient decent (*SGD* in torch.optim) with a learning rate of 0.001 and momentum of 0.9 was used as the optimizer. Data was loaded into the CNN in batches of 32 with shuffling (*DataLoader* in torch.utils.data), and backwards propagation and parameter optimization were conducted in such batches. Models were trained for 10 epochs, and model parameters from the epoch with the best accuracy on the validation set were chosen as the final model parameters. The model was then tested on the strides of the left-out participant. Model predictions for whether each stride from the left-out participant was from a control or an OA participant were aggregated across the *N* CNN models, and the overall classification performance was computed.

### CNN model for subject classification and latent representation

As the digital insoles produced high-frequency raw sensor time-series data, we analyzed whether such structured strides (50 measurements along 100 interpolated timepoints for each stride) contained informative subject-specific gait features. To utilize the temporal aspect of the data, we constructed a one-dimensional CNN in which the model could interpret the relationship between sequential timepoints for each sensor. This temporal relationship in the input data was lost in our previous analysis, in which the stride was flattened and interpreted by XGBoost.

For the individuality and consistency CNN models, the model was trained to identify the subject from which a stride came. However, the purpose of using the CNN model was not to classify training subjects based on their strides, but rather to extract activation of the penultimate fully connected layer for the model’s latent representation of the “ gait fingerprint” of a stride. As such, once the CNN model was trained on participants in the training set, the model was then applied to participants in the hold-out testing set and latent representations for each stride were extracted.

To train the CNN model, strides from the training participants were split into a 64% training set, a 16% validation set, and a 20% final validation set. A similar CNN architecture was used as before, except now rather than a binarized control versus OA output, the model outputs the subject label. As such, the model architecture differed starting from the second fully connected layer:

- Second fully connected layer with 120 in channels and 60 out channels.
- Element-wise rectified linear activation unit.
- Dropout with 0.2 probability.
- Third fully connected layer with 60 in channels and 23 out channels.

The CNN model was trained in the same manner as before, except multi-class cross entropy loss (*CrossEntropyLoss*) was used as the loss function. As before, the model was trained for 10 epochs, and model parameters from the epoch with the best accuracy on the validation set were chosen as the final model parameters. The final validation set was then used to check the final model’s performance. A forward hook (*register_forward_hook* in torch.nn.modules) was attached to the penultimate fully connected layer, to extract activation of that 60-element layer for a new stride inputted into the model.

### Evaluation of subject individuality across different representations

Models were constructed for each data type to predict individual subjects in the training set, and then applied on the testing set. Next, distances between each pair of walks/strides were calculated within a subject, within other subjects with the same disease status, and within other subjects with a different disease status.

Each feature was first z-scored (centered and scaled to unit variance, using *scale* function in base R), and Euclidean distances between all walks/strides in the testing set were calculated using *dist* function in the R stats package. To compare across representations with differing number of features, distances were divided by the square root of the number of features. The mean distance between every two participants (including with oneself) was then calculated.

To evaluate models for subject individuality, each participant-to-participant comparison was categorized into the groups of control within-self, OA within-self, control with another control, OA with another OA, or one control with one OA. Significance of difference in distances between participant categories was analyzed with t-tests in the R stats package. Effect sizes as Cohen’s D were computed with the R effsize package using default settings. For all boxplots shown, the center line indicates the median, and the box limits indicate the upper and lower quartiles.

### Evaluation of CNN models of subject individuality and consistency

Digital insole sensor data from both baseline (day 1) and on-treatment timepoints (day 85) of OA participants in the R5069-OA-1849 clinical trial was used to evaluate whether training on data from two days instead of just one day improves the consistency of the CNN representation of participants. A second consistency CNN model was trained on combined data from both timepoints for training set participants, where input data was labeled only by participant identity and not by timepoint. For comparability, both the individuality and consistency CNN models used the same split of train and test participants. Both models were given day 1 and day 85 of testing set participants, and the distance between all stride pairs as represented by the penultimate CNN layer was calculated as before.

To evaluate models for consistency, each participant-to-participant comparison was then categorized into the groups of within-self same day, within-self different day, or subject with another subject. Only OA participants were analyzed for consistency as only they were assessed on two different days. Significance of difference in distances between the CNN individuality and consistency models across the same participant comparisons was analyzed with paired t-tests in the R stats package. For all boxplots shown, the center line indicates the median, and the box limits indicate the upper and lower quartiles.

## Supplementary Figures and Tables

**Supplementary Fig. 1.**
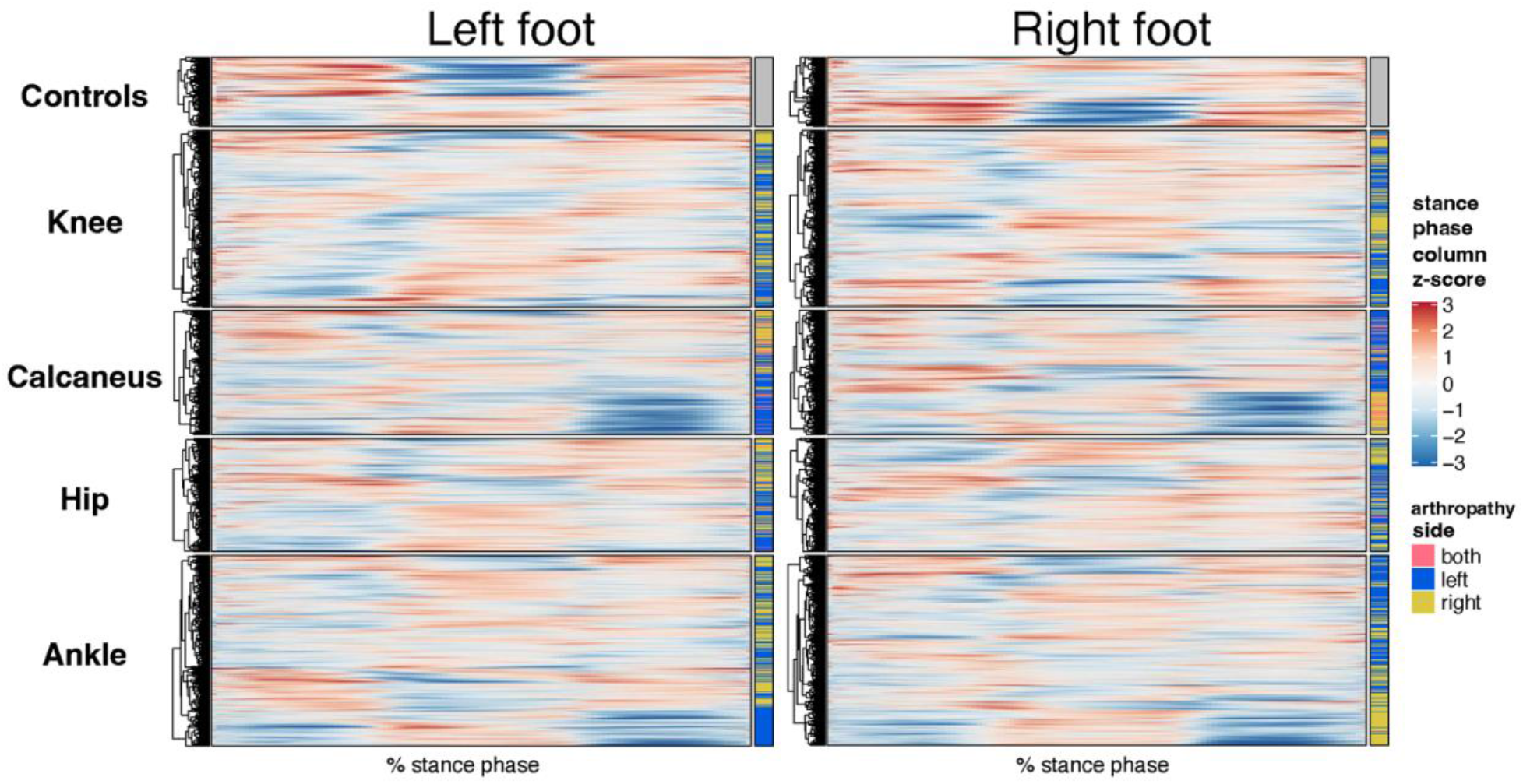
Heatmap representation of vertical ground reaction force (vGRF) data from GaitRec dataset for all joints with injuries and controls. ^**25**^ Data are z-scored by each column (% stance phase) across all walks. Heatmaps are separate by injury class (control, knee, calcaneus, hip, and ankle), and vGRF from each walk are unsupervised clustered within each category. The right of the heatmap annotates the joint side with the arthropathy (left joint, right joint, both joints, or no injury in the control group).

**Supplementary Figure 2.**
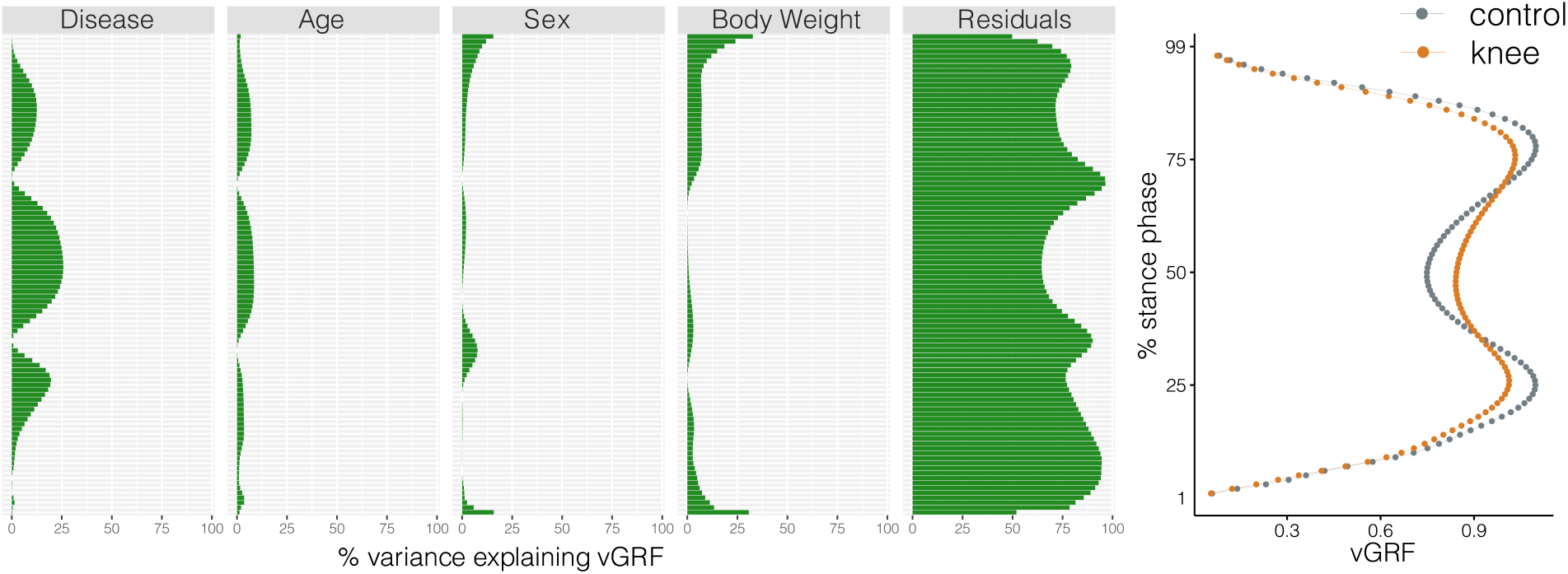
Variance explained in vertical ground reaction force (vGRF) with clinical and demographic characteristics of the participants. Linear models were fit at each % stance phase (timepoint), excluding the edges of the curve which are bounded by 0 (and as such have no variance). We used disease (knee arthropathy or control), age, sex (male or female), and body weight as covariates in the model, with each subsequent vGRF % stance phase timepoint as the dependent variable. Within each linear model, using the sum of squares for each category compared to the total sum of squares, we calculated of the variance each component’s contribution to the total variance, with the residuals indicating the unexplained variance in these models. We observed that the disease state is the major contributor to vGRF for most of the curve, with age, sex, and body weight also explaining a smaller proportion of the variance.

**Supplementary Fig. 3.**
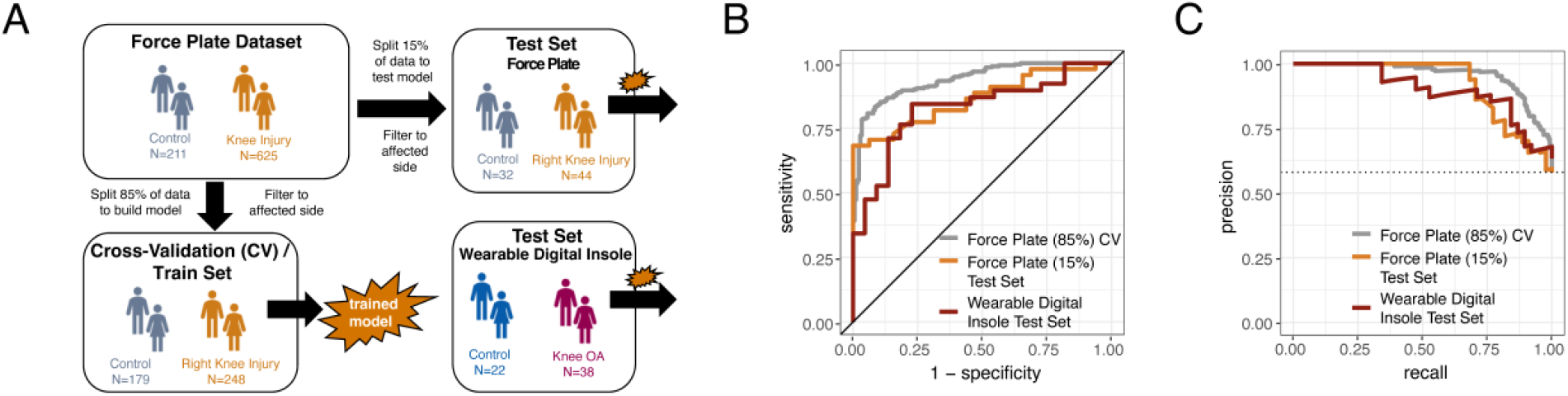
**a**, Schematic of machine learning model building of training/validation and testing sets with the right foot data, as in Fig. 2. b, Receiver operating characteristic curve for XGBoost classification of force plate (85%) cross-validation (CV, training/validation) set, force plate (15%) hold-out test set, and the digital insole test set for right foot data. c, Precision-recall curve for XGBoost classification of the same groups in b for right foot data.

**Supplementary Fig. 4.**
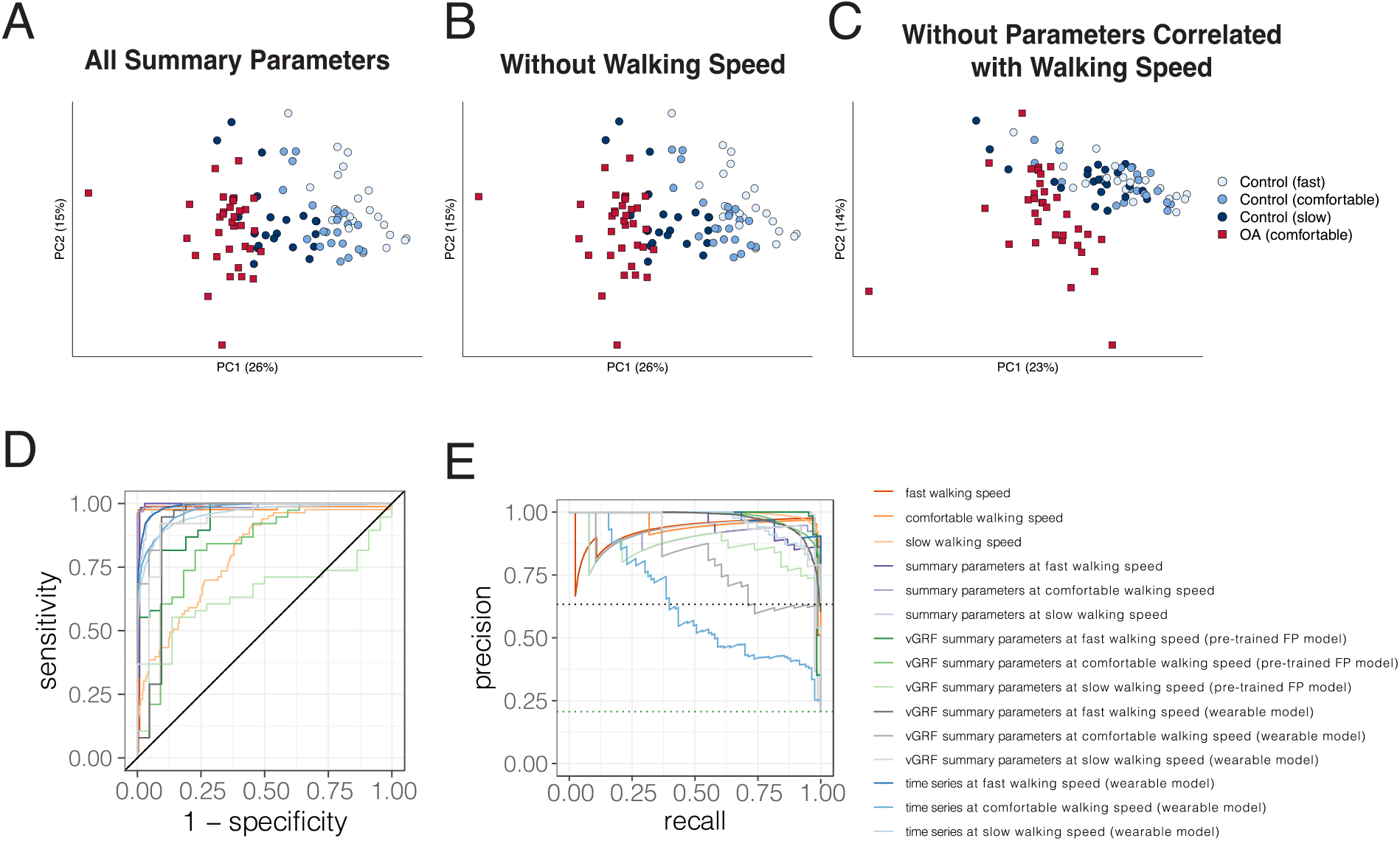
**a**, Principal component analysis (PCA) of all derived gait characteristics measured using the Moticon insole device, where each point represents the average of all walks from a particular subject, and the dot color indicates the group (control or knee osteoarthritis [OA]) or walking speed of control subjects. b, PCA analysis as in a, without the walking speed gait characteristic. c, PCA analysis as in a, without the 14 derived gait characteristics correlated to walking speed. d, Classification performance auROC using walking speed as a sole predictor, vertical ground reaction force (vGRF) data, derived gait characteristics, and time series data.

**Supplementary Fig. 5.**
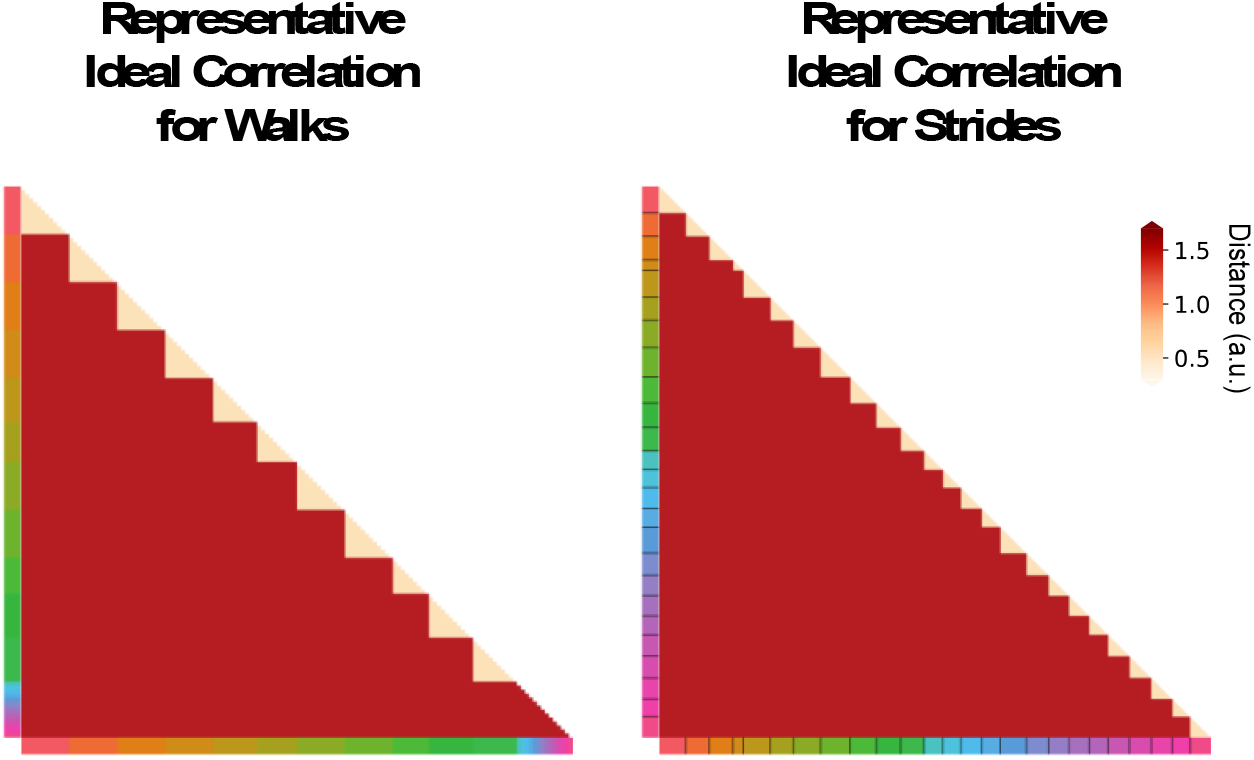
Example heatmap of a good representation that has low distance between all pairs of walks/strides from the same participant and high distance between all pairs of walks/strides from different participants. Color along the edge indicates each person.

**Supplementary Fig. 6.**
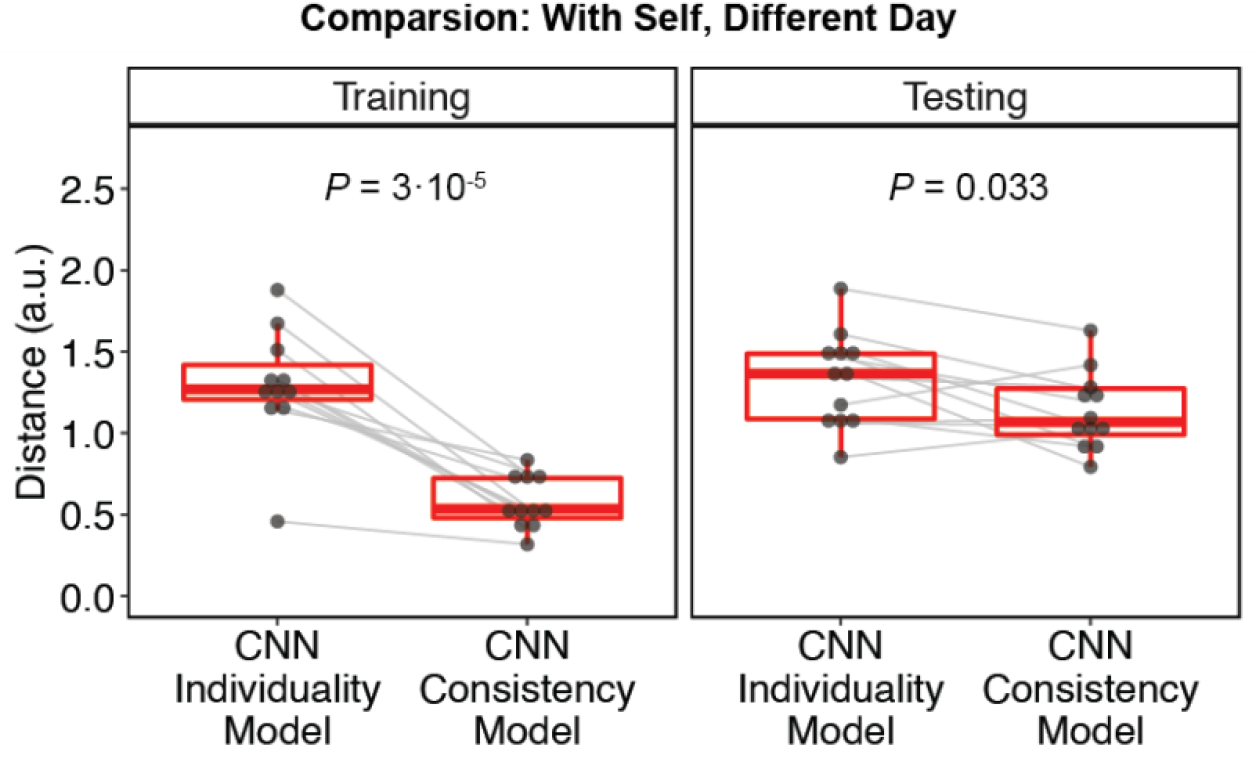
Boxplots of mean distance (in arbitrary units) of each stride with other strides from the same person on different days, for both the convolutional neural network (CNN) individuality model (Fig. 5) and CNN consistency model (Fig. 6a,b) in both the training and testing sets. Values are replotted from Fig. 6c, and lines are drawn between the same participants. Significance of difference in distances between the CNN individuality and consistency models was analyzed with two-sided paired t-tests.

**Supplementary Fig. 7.**
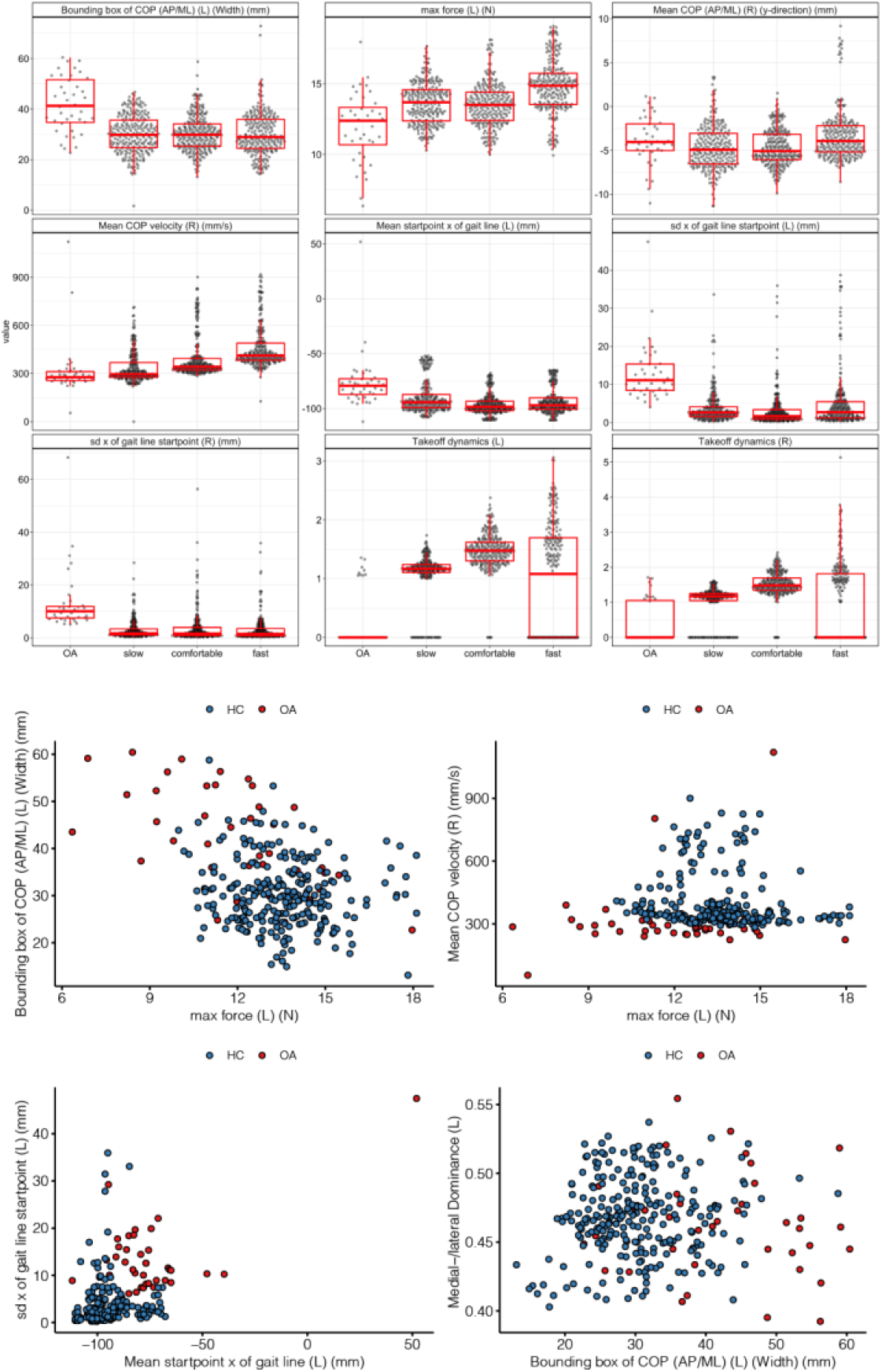
Derived gait characteristics that are most discriminative of knee osteoarthritis (OA) versus controls include features shown in Supplementary Table 1. **Top**, Boxplots in knee OA, control slow, comfortable, and fast walking speeds for some parameters predictive of knee OA versus controls. **Bottom**, Scatter plots of select parameters in HC (healthy control) versus OA at comfortable walking speed.

**Supplementary Table 1.**
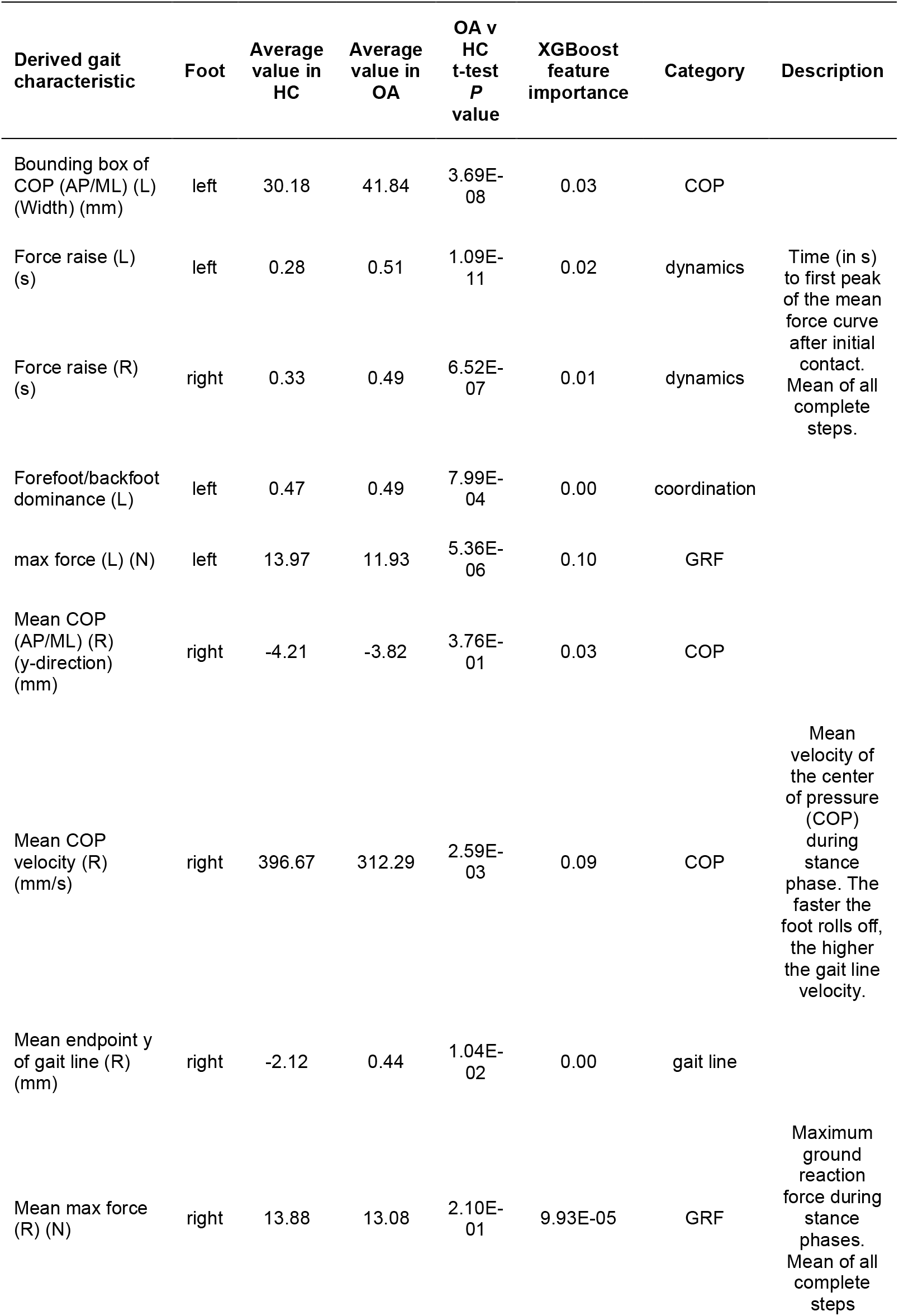
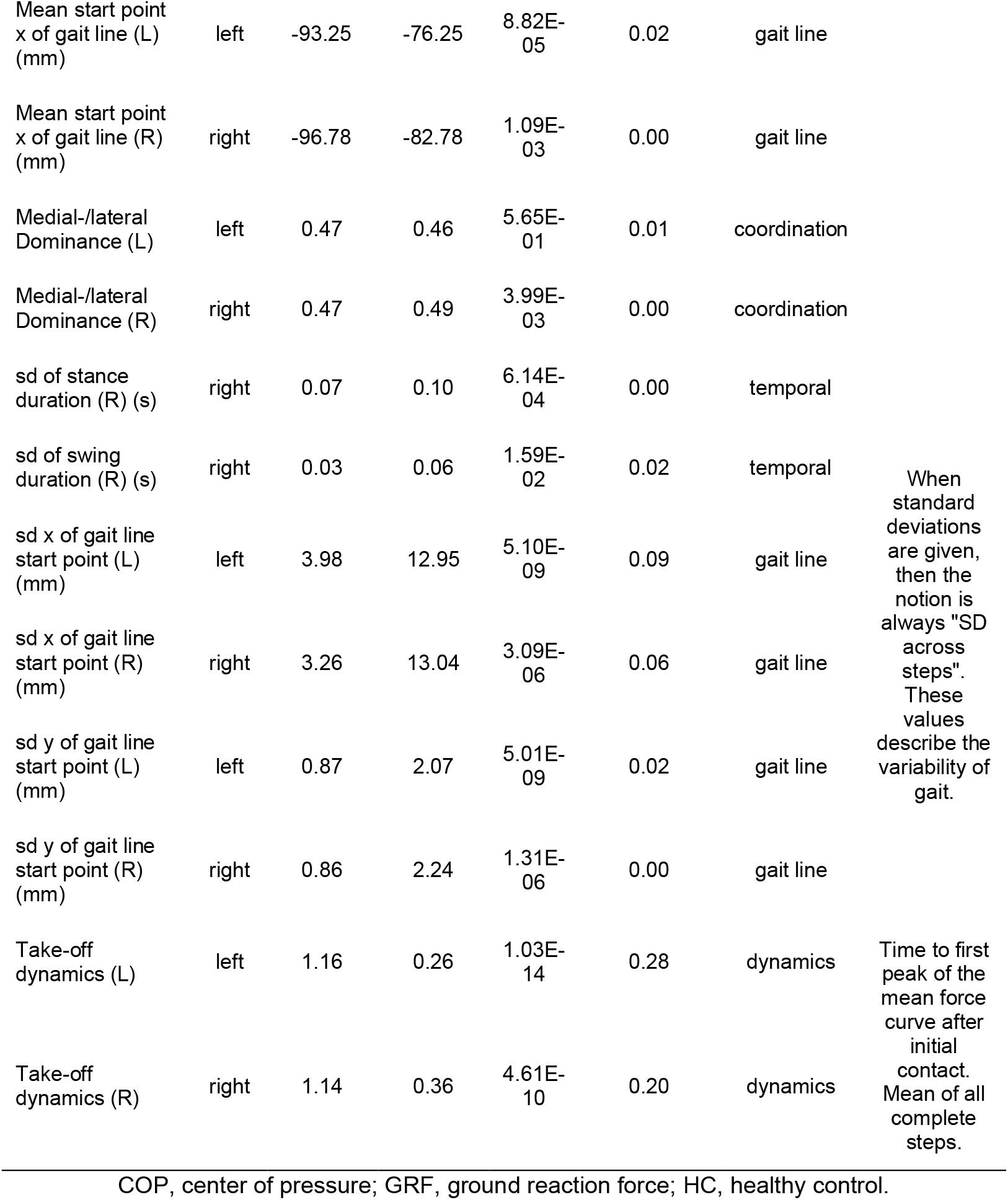
Derived gait characteristics associated found to be important in the machine learning (ML) model to differentiate control from knee osteoarthritis (OA) subjects.

## Notes

### Clinical Trial

NCT03956550

### Clinical Protocols

https://clinicaltrials.gov/ProvidedDocs/50/NCT03956550/Prot_000.pdf

### Author Declarations

The study protocol received Institutional Review Board and ethics committee approvals from Moldova Medicines and Medical Device Agency and National Ethics Committee for Moldova, and the Western Institutional Review Board.

